# Increased infections, but not viral burden, with a new SARS-CoV-2 variant

**DOI:** 10.1101/2021.01.13.21249721

**Authors:** A. Sarah Walker, Karina-Doris Vihta, Owen Gethings, Emma Pritchard, Joel Jones, Thomas House, Iain Bell, John I Bell, John N Newton, Jeremy Farrar, Ian Diamond, Ruth Studley, Emma Rourke, Jodie Hay, Susan Hopkins, Derrick Crook, Tim Peto, Philippa C. Matthews, David W. Eyre, Nicole Stoesser, Koen B. Pouwels, COVID-19 Infection Survey team

**Author notes:** Corresponding author: A. Sarah Walker.

## Abstract

**Background:** A new variant of SARS-CoV-2, B.1.1.7/VOC202012/01, was identified in the UK in December-2020. Direct estimates of its potential to enhance transmission are limited.

**Methods:** Nose and throat swabs from 28-September-2020 to 2-January-2021 in the UK’s nationally representative surveillance study were tested by RT-PCR for three genes (N, S and ORF1ab). Those positive only on ORF1ab+N, S-gene target failures (SGTF), are compatible with B.1.1.7/VOC202012/01. We investigated cycle threshold (Ct) values (a proxy for viral load), percentage of positives, population positivity and growth rates in SGTF vs non-SGTF positives.

**Results:** 15,166(0.98%) of 1,553,687 swabs were PCR-positive, 8,545(56%) with three genes detected and 3,531(23%) SGTF. SGTF comprised an increasing, and triple-gene positives a decreasing, percentage of infections from late-November in most UK regions/countries, e.g. from 15% to 38% to 81% over 1.5 months in London. SGTF Ct values correspondingly declined substantially to similar levels to triple-gene positives. Population-level SGTF positivity remained low (<0.25%) in all regions/countries until late-November, when marked increases with and without self-reported symptoms occurred in southern England (to 1.5-3%), despite stable rates of non-SGTF cases. SGTF positivity rates increased on average 6% more rapidly than rates of non-SGTF positives (95% CI 4-9%) supporting addition rather than replacement with B.1.1.7/VOC202012/01. Excess growth rates for SGTF vs non-SGTF positives were similar in those up to high school age (5% (1-8%)) and older individuals (6% (4-9%)).

**Conclusions:** Direct population-representative estimates show that the B.1.1.7/VOC202012/01 SARS-CoV-2 variant leads to higher infection rates, but does not seem particularly adapted to any age group.

## INTRODUCTION

After initial reductions in SARS-CoV-2 cases, infection rates have increased in many countries following release of large-scale lockdowns^1^. There are likely multiple reasons, including variable adherence to different levels of restrictions. However, the potential contribution of genetic adaptation has been highlighted by the discovery of a new genetic variant in southern England, B.1.1.7/VOC(variant of concern)202012/01^2^, followed by rapid increases in cases. Amongst unusually large numbers of genetic changes supporting this lineage, particularly in the spike (S) protein, three have potentially important effects. N501Y in the receptor binding domain increases binding affinity to the ACE2 receptor^3,4^, H69del/V70del may be associated with immune response evasion, and P681H, adjacent to the furin cleavage site^5^, may facilitate entry into epithelial cells. H69del/V70del also affects PCR assays targeting the S-gene, preventing probe binding and causing S-gene target failure (SGTF).

To date, data on the transmissibility and severity of B.1.1.7/VOC202012/01 have come from two mathematical modelling studies (using hospital admissions/occupancy, deaths, prevalence [from symptomatic community testing] and seroprevalence, and percentages of sequenced positives accounted for by B.1.1.7/VOC202012/01 in southern England)^6,7^, and analyses of the UK’s symptomatic community testing programme^7-9^ and whole genome sequences from this programme^7,9^. Estimates suggest 56%^6^ (95% credible interval 50-74%) and 40-80%^7^ greater transmissibility associated with B.1.1.7/VOC202012/01. Analyses of community testing are limited by non-random sampling of symptomatic individuals, and there being only three laboratories in the UK’s national testing programme which can identify SGTFs^10^, meaning only ∼35% tests can be categorised,^7^ distributed unequally nationally. However, at present SGTF is highly sensitive (99.3%) and specific (99.5%) for H69del/V70del^8^, with the percentage of H69del/V70del that were B.1.1.7/VOC202012/01 increasing from 3% mid-October to 64% early-November and 98% early-December^11^. A matched case-control study of 3,538 fully sequenced B.1.1.7/VOC202012/01 and wild-type cases found no evidence of differences in hospitalisation (0.9% vs 1.5%, respectively) or 28-day mortality (0.9% vs 0.7%, respectively)^8^. However, secondary attack rates were raised in contacts of B.1.1.7/VOC202012/01 (14.7%) or SGTF (14.9%) versus wild-type (11%)^11^.

Here we use the UK’s national COVID-19 Infection Survey (CIS), a representative sample of households with longitudinal follow-up^12^, in which SGTF is consistently identified, to directly investigate whether the new variant is associated with higher infection rates, overall or particularly in children, given that schools generally remained open despite other lockdown precautions during the study period.

## METHODS

This analysis included all SARS-CoV-2 RT-PCR tests of nose and throat swabs from 28-September-2020 to 2-January-2021 in the Office for National Statistics (ONS) CIS (ISRCTN21086382, https://www.ndm.ox.ac.uk/covid-19/covid-19-infection-survey/protocol-and-information-sheets). The survey randomly selects private households on a continuous basis from address lists and previous surveys to provide a representative UK sample. Following verbal agreement to participate, a study worker visited each household to take written informed consent, which was obtained from parents/carers for those 2-15 years; those aged 10-15 years provided written assent. Those <2 years were not eligible.

Individuals were asked about demographics, symptoms, contacts and relevant behaviours (https://www.ndm.ox.ac.uk/covid-19/covid-19-infection-survey/case-record-forms). To reduce transmission risks, participants ≥12 years self-collected nose and throat swabs following study worker instructions. Parents/carers took swabs from children <12 years. At the first visit, participants were asked for (optional) consent for follow-up visits every week for the next month, then monthly for 12 months from enrolment. The study received ethical approval from the South Central Berkshire B Research Ethics Committee (20/SC/0195).

Swabs were analysed at the UK’s national Lighthouse Laboratories at Milton Keynes and Glasgow using identical methodology. RT-PCR for three SARS-CoV-2 genes (N protein, S protein and ORF1ab) used the Thermo Fisher TaqPath RT-PCR COVID-19 Kit, and analysed using UgenTec FastFinder 3.300.5, with an assay-specific algorithm and decision mechanism that allows conversion of amplification assay raw data from the ABI 7500 Fast into test results with minimal manual intervention. Samples are called positive if at least a single N-gene and/or ORF1ab are detected (although S-gene cycle threshold (Ct) values are determined, S-gene detection alone is not considered sufficient to call a sample positive). We estimated a single Ct value as the arithmetic mean of Ct values for genes detected (Spearman correlation >0.98 between each pair of Ct values). Viral loads were estimated from a linearity curve (in^13^, Supplementary Figure 1).

**Figure 1.**
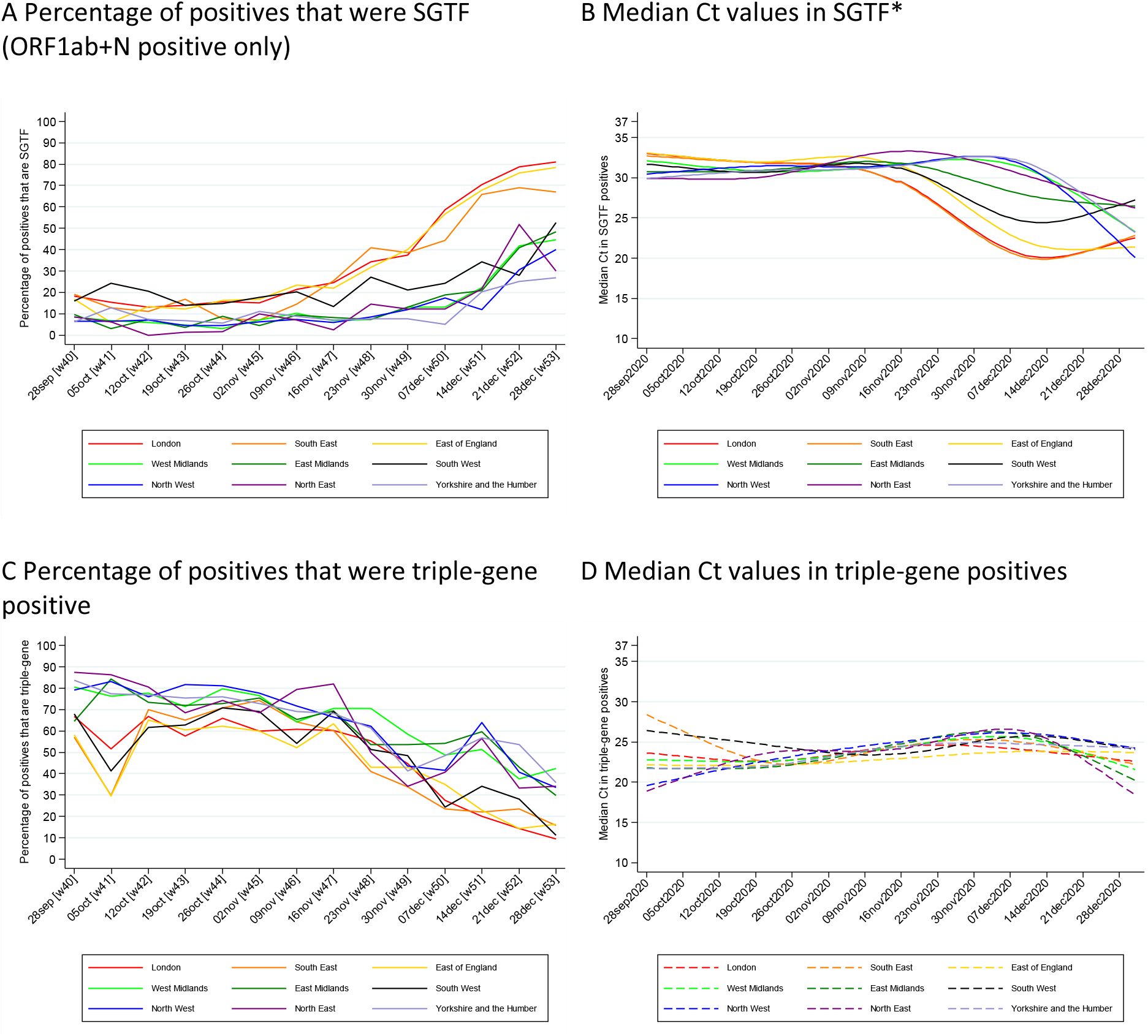
Percentage of positives and Ct values over time, English regions. * whole genome sequencing of community symptomatic testing samples in England showed that 3% samples with H69del/V70del were B.1.1.7/VOC(variant of concern)202012/01 mid-October (N=116), rising to 64% at the start of November (N=398), 88% mid-November (N=602) and 98% at the start of December (N=2,007)^1^. Note: Ct 30 ∼150 copies/ml, 25 ∼5500 copies/ml, 20 ∼230,000 copies/ml based on linearity curves (in^2^ Supplementary Figure 1). Devolved Administrations shown in **Supplementary Figure 1**.

The presence of 12 specific symptoms in the previous seven days was elicited at each visit (cough, fever, myalgia, fatigue, sore throat, shortness of breath, headache, nausea, abdominal pain, diarrhoea, loss of taste, loss of smell), as was whether participants thought they had (unspecified) symptoms compatible with COVID-19. Any positive response to any symptom question at the swab-positive visit defined the case as symptomatic at the test.

We investigated Ct values using median (quantile) regression, positivity rates at the regional level using multi-level regression and post-stratification (MRP), and growth rates using iterative sequential Poisson regression (ISR)^14,15^ (details in Supplementary Methods). Cases positive only on a single gene consistently have high Ct values reflecting variable target detection at low viral loads^13^. Therefore, we considered only samples positive for ORF1ab+N-gene and negative for S-gene as SGTF, not those positive for the N-gene or ORF1ab alone. Sensitivity analyses included only positives with Ct<30, to assess the impact of non-SGTF cases being more likely to be sampled late in their infection, and hence have higher Ct values^16^.

## RESULTS

From 28-September-2020 to 2-January-2021, 372,626 participants from 185,342 households across the United Kingdom had results from median 4 (IQR 3-5, range 1-12) nose and throat swabs each. Of 1,553,687 test results, 15,166 (0.98%, 95% CI 0.96-0.99%) were positive, in 11,800 individuals from 8,868 households. 8,545(56%) were positive on all three genes, 3,531(23%) only on ORF1ab+N, i.e. were SGTF compatible with B.1.1.7/VOC202012/01, and 3,090(20%) were other single/double positives.

SGTF comprised an increasing, and triple-gene positives a decreasing, percentage of positives from late-November in most regions/countries, consistent with SGTF representing non-B.1.1.7/VOC202012/01 H69del/V70del strains, or positives with lower viral load, before this (Figure 1A/C, Supplementary Figure 1). The timing of rises in SGTF-positives varied strongly across region/country; e.g., rising from 15% early-November to 38% end-November and 81% end-December in London, versus 7%, 13% and 45% respectively in the West Midlands. Results were similar restricting to positives with Ct<30, which may be more likely to be new infections (Supplementary Figure 2). In parallel, Ct values in SGTF showed a major shift, being consistently high (∼30, ∼150 copies/ml) through to mid-November, before dropping sharply to a minimum ∼20 (∼230,000 copies/ml) at different times depending on region/country (Figure 1B), likely reflecting expansion of B.1.1.7/VOC202012/01 amongst SGTF positives^8^. In contrast, Ct values varied much less in triple-gene positives, remaining ∼22-27 (Figure 1D), with small increases and decreases consistent with trends in overall positivity^13,16^. By January 2021, median Ct values were similar in SGTF and triple-gene positives in most regions/countries, suggesting that earlier differences were due to more SGTF being new infections rather than intrinsic biological differences affecting viral loads.

**Figure 2.**
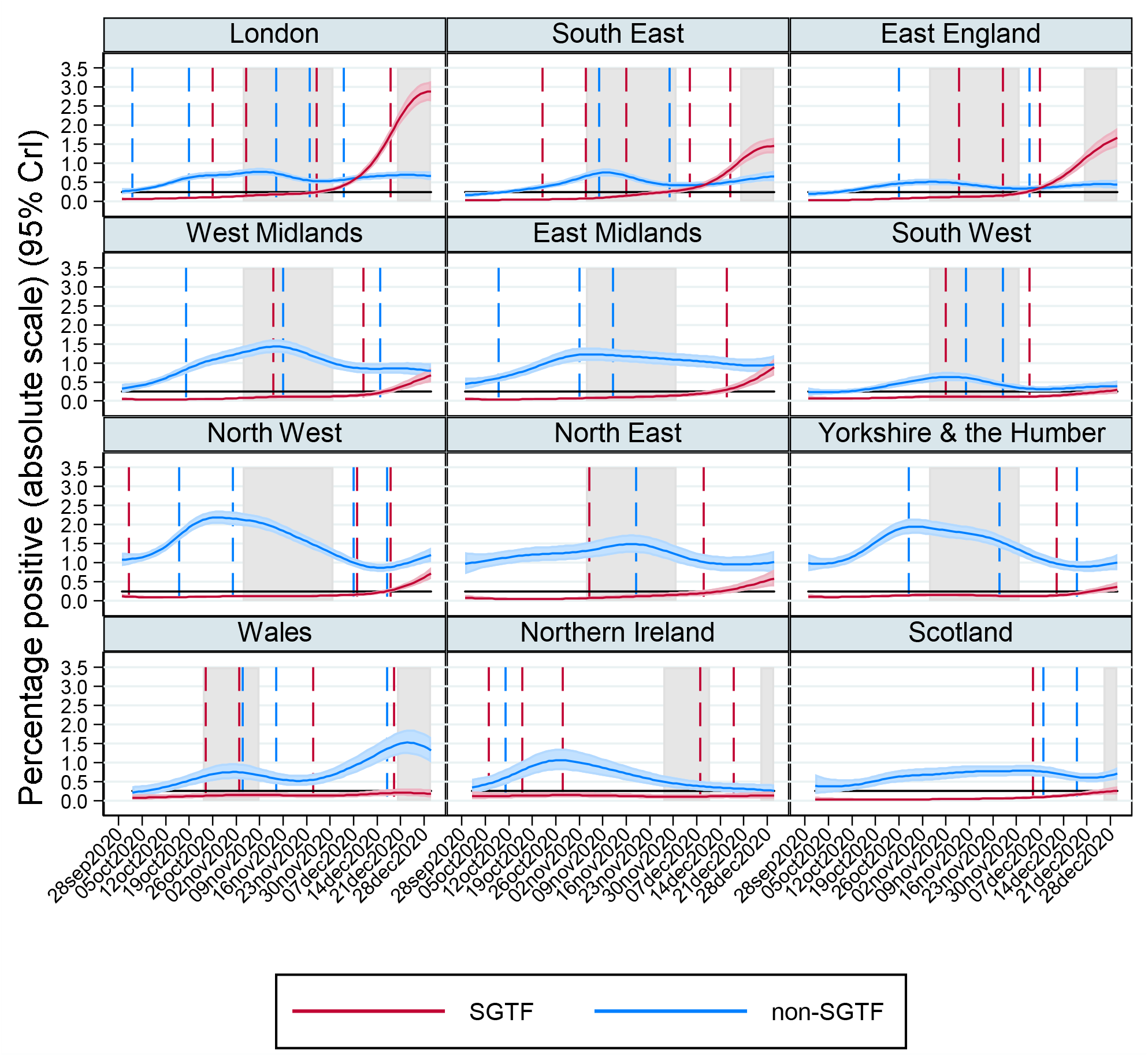
Percentage of the population positive with SGTF (ORF1ab+N positive, compatible with B.1.1.7/VOC202012/01) and non-SGTF. Note: gray shading shows national restrictions/stay at home orders for the majority of the region. Black horizontal line at 0.25%. Dashed lines show estimated changes in trend from ISR algorithm fitted from 1 Sept (no dashed line means no change in trend with p<0.01 (non-SGTF) or p<0.05 (SGTF) detected). See **Supplementary Figure 3** for probabilities on the log scale that is used for ISR modelling.

At a population level, the percentage of individuals with SGTF vs non-SGTF positives in different regions/countries varied substantially over time (Figure 2, Supplementary Figure 3). Marked increases in SGTF positivity rates occurred in London, the South East and East of England from late-November despite rates of non-SGTF positives remaining stable, suggesting trends were not due to changing behaviour alone. In more northern English regions, increases in SGTF positivity started later, from mid-December, generally on a background of stable rates of non-SGTF positives. SGTF positivity remained relatively low in the South West, Wales, Northern Ireland, and Scotland, furthest from the South East. Analyses considering household positivity were similar (Supplementary Figure 4).

**Figure 3.**
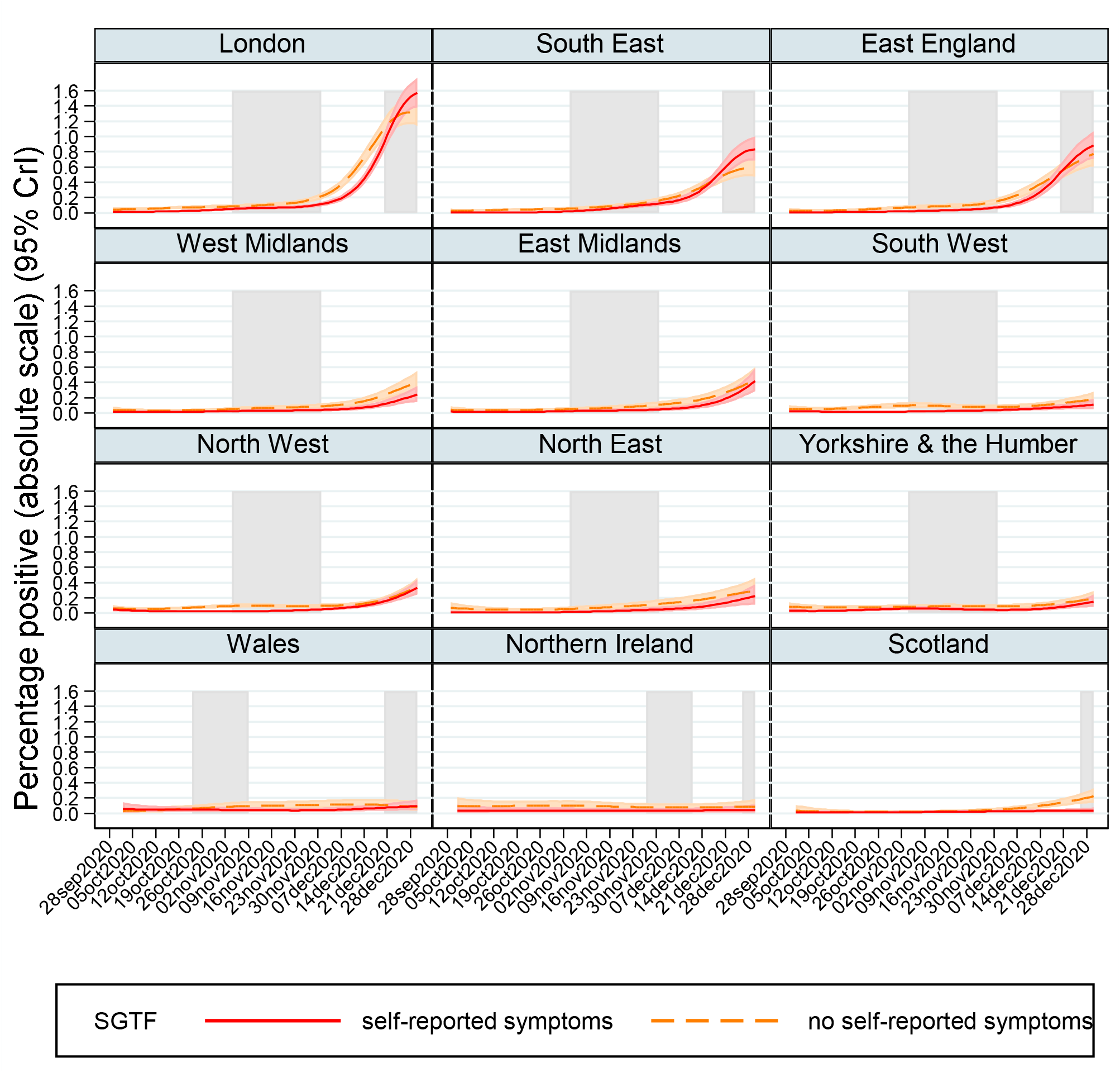
Percentage of the population positive with SGTF (ORF1ab+N positive, compatible with B.1.1.7/VOC202012/01) according to self-reported symptoms at the test. Note: gray shading shows national restrictions/stay at home for the majority of the region.

**Figure 4.**
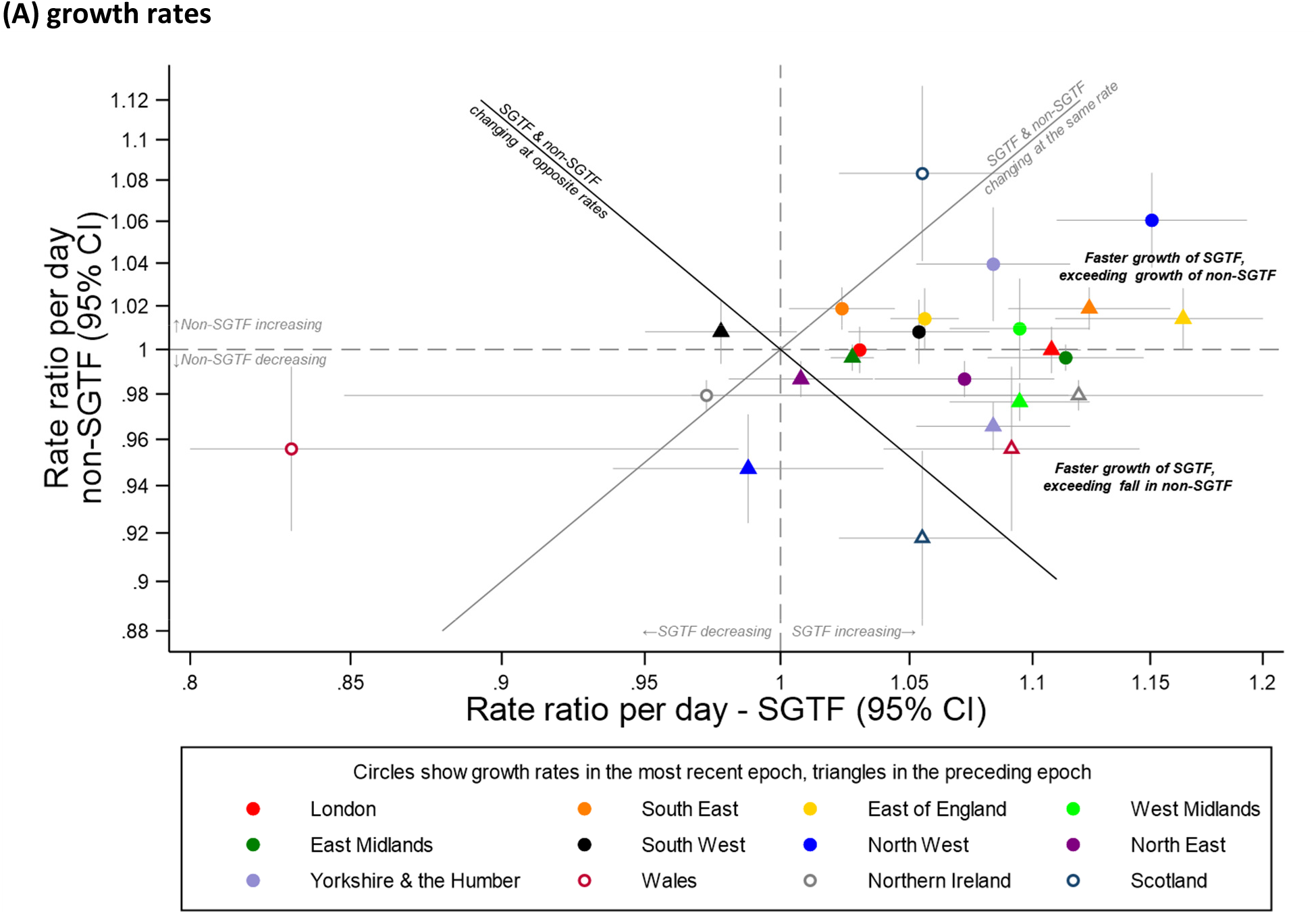
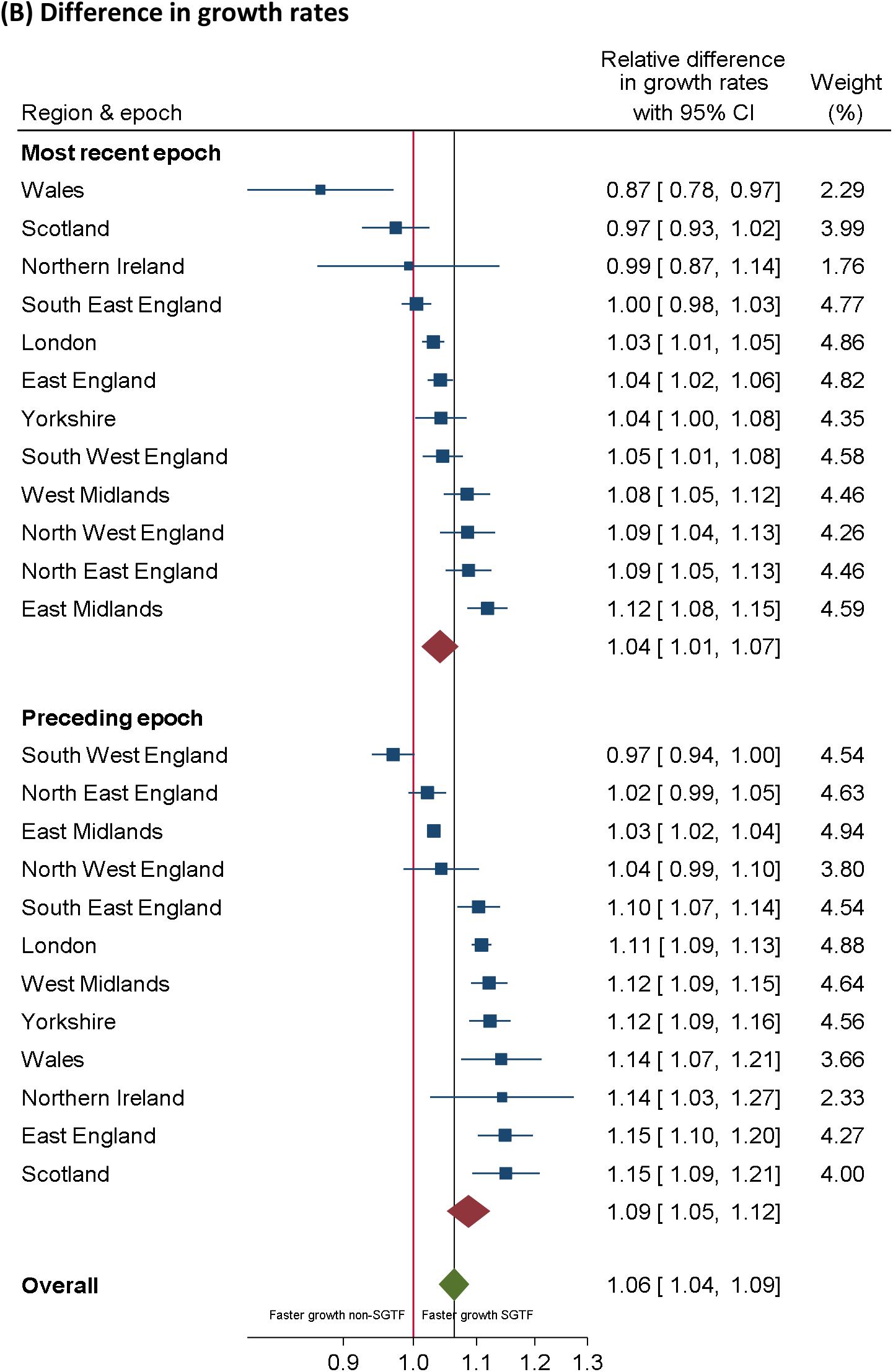
Growth rates of SGTF and non-SGTF positives in two most recent epochs defined by ISR. Note: panel (A) shows growth rates (rate ratio (RR) per day) of SGTF (x-axis) and non-SGTF (y-axis) positives within the same region in epochs defined by changepoints (change in trend) identified by ISR and shown in **Figure 2** and **Supplementary Table 1**. 95% CI are truncated at 0.8 and 1.2. RR>1 mean positivity rates are increasing, <1 that they are decreasing. Points to the right of the gray diagonal line are periods of time where, within one region, SGTF positives are increasing faster than non-SGTF positives; and points on/around the gray line where SGTF and non-positives are changing at similar rates within a region. The black diagonal line indicates opposite growth rates, that is SGTF are increasing at the same rate non-SGTF are decreasing or vice versa within a region, consistent with replacement. Points above the black line are consistent with addition. Panel B shows difference between growth rates in SGTF and non-SGTF from **Figure 4A**, combined using random effects meta-analysis.

Importantly, increases in SGTF positivity were similar in those with and without self-reported symptoms (Figure 3). Trends over time in non-SGTF positivity were similar in those with and without self-reported symptoms (Supplementary Figure 5), but rates were generally slightly higher in those not reporting symptoms, potentially reflecting ongoing PCR-positivity after earlier acute infection. Supporting this, rates of non-SGTF positivity with Ct<30 (i.e. more likely early infection) were much more similar in those with and without self-reported symptoms (Supplementary Figure 6).

**Figure 5.**
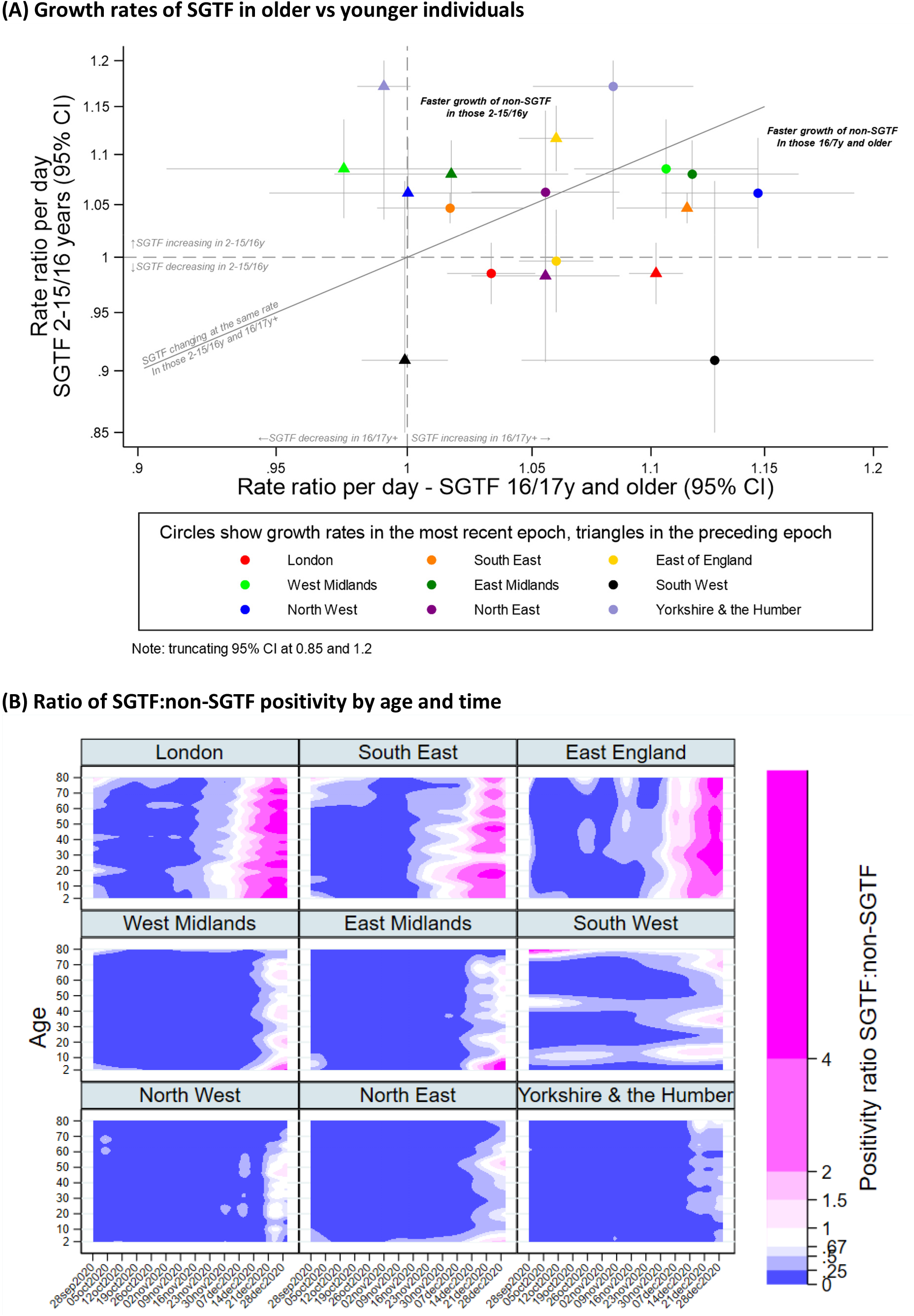
Impact of age on SGTF growth rates (A) and ratio of SGTF:non-SGTF positivity. Note: for panel A, meta-analysis combining estimates shown in **Supplementary Figure 12B**, and corresponding estimates for non-SGTF in **Supplementary Figure 12A&C**. SGTF and non-SGTF positivity rates by age over time shown in **Supplementary Figure 15**.

The relative dynamics of SGTF vs non-SGTF varied over time (Figure 2). The most marked increases in growth rates for SGTF occurred at a median positivity rate of 0.25% (range 0.15-1%, Supplementary Figure 7). To assess whether B.1.1.7/VOC202012/01 was replacing or adding to existing strains, we compared growth rates of SGTF vs non-SGTF within the same region in the two most recent epochs defined by changes in trend in either SGTF/non-SGTF identified by ISR (Figures 2&4A, Supplementary Table 1, Supplementary Figure 8). SGTF positivity rates increased more rapidly than non-SGTF positivity rates in most regions in most epochs (points to the right of the gray line in Figure 4A), but particularly strongly in the preceding epoch in southern regions and the most recent epoch in East/West Midlands, the North West and Yorkshire and the Humber (doubling times all under 10 days, Supplementary Table 1). Further, the rate of growth in SGTF positives generally exceeded the decline in non-SGTF positives (points above the black line), by an average of 6% (95% CI 4-9%) (Figure 4B), supporting addition rather than replacement with the B.1.1.7/VOC202012/01. In January 2021,

SGTF growth rates were stabilising in the regions with the highest absolute rates, but increasingly markedly in regions with intermediate rates such as the Midlands and North West (Supplementary Figure 9A). Sensitivity analyses restricting to “new infections” with Ct<30 were similar (Supplementary Figure 10).

The relative difference in growth rates of SGTF vs non-SGTF had a similar distribution in those up to high-school age (i.e. ≤15/16 years, 5% excess (95% CI 1-8%)) versus older (6% (4-9%)) (Supplementary Figure 11, Supplementary Table 2), with no evidence that SGTF positivity rates were consistently growing faster or slower in those under and over high school age (Figure 5A; similarly for non-SGTF positives, Supplementary Figure 12A). Consistent with this, there was no evidence of variation by age group in the percentage of positives that were SGTF vs non-SGTF between 21-December-2020 and 2-January-2021 for any region/country (p>0.05, Supplementary Figure 13, Supplementary Table 3). However, because positivity was generally highest in those up to high school age (Supplementary Figure 14), a greater number of infections would be expected to arise in younger individuals from the same growth rate as in older individuals.

Lastly, we estimated the ratio of SGTF positivity rates by age (Supplementary Figure 15A) to non-SGTF positivity rates by age (Supplementary Figure 15B) in the nine English regions with sufficient cases to model age as a continuous factor. Although increases in SGTF positives relative to non-SGTF positives generally occurred earliest in younger individuals, this was not uniform across regions and did not occur consistently in school-age children first (Figure 5B).

## DISCUSSION

In this large representative community surveillance study, we found clear evidence of increases in SGTF SARS-CoV-2, consistent with B.1.1.7/VOC202012/01 expansion, in multiple UK regions from mid-to-late November, including during periods of national lockdown when non-SGTF strains were stable or decreasing. Multiple lines of evidence support B.1.1.7/VOC202012/01 leading to higher infection rates in adults and children, and adding to, rather than simply replacing, existing strains. However, we found no evidence that Ct values (a proxy for viral load) were intrinsically substantially lower in SGTF-positives, in contrast to initial reports^17,18^, but consistent with observations that B.1.1.7/VOC202012/01 infection is not more severe^8^. Importantly, rates of SGTF infections with and without self-reported symptoms were similar, consistent with the higher prevalence of asymptomatic infection reported in defined populations (30%^19^) and community surveillance (e.g. 42%^20^, 72%^21^). Asymptomatic infections may therefore be contributing substantially to B.1.1.7/VOC202012/01 spread, and are not currently captured by the national testing programme, which focusses on symptomatic cases and their contacts.

Genetic drift means that new variants will continuously arise, and some will become the established dominant viral population simply due to chance founder effects. Here we directly compared growth rates for SGTF vs non-SGTF strains within epochs within regions (i.e. matched) to attempt to control for variation in behaviour and other confounders which undoubtedly affect transmission. SGTF consistently had greater growth rates, supporting increased transmissibility, since it is unlikely that chance founder effects would have the same influence in multiple regions/countries. Further, excess SGTF growth rates generally outweighed declines in non-SGTF positives, showing B.1.1.7/VOC202012/01 is likely adding to, rather than replacing, existing strains. This is particularly concerning given recent reports of the recent expansion of SGTF in the US^22^, although this may reflect different variants^23^. Our findings of no evidence of difference in SGTF growth rates between children and adults do not support B.1.1.7/VOC202012/01 being particularly adapted to transmit more in children. However, the higher current positivity rates in children and young adults in the UK inevitably means that the same growth rates will result in a higher number of new infections from these age groups.

The marked increases in SGTF varied substantially in timing between regions. B.1.1.7/VOC202012/01 was first observed end-September^2^; our analyses are consistent with its presence at low but increasing frequencies in southern regions from this time (Supplementary Figure 3). Increases generally became marked once 0.25% SGTF positivity was exceeded (Figure 2); heterogeneity in dispersion (“k”) and super-spreading events, particularly from those without symptoms but with low Ct/high viral loads^24^, plus chance variation, could explain its persistence without inevitable growth below this.

We found no evidence that increased infection rates associated with B.1.1.7/VOC202012/0 were mediated through higher viral load (lower Ct). Direct comparisons are complicated by the fact that average Ct values in surveillance studies depend on whether positivity rates are increasing or decreasing^16^; however, current medians in SGTF are similar to those observed in triple-positives in regions with high positivity rates (e.g. northern England in early October). Further, sensitivity analyses restricting to positives with Ct<30, more likely to be new infections rather than ongoing PCR-positivity, gave similar results. An alternative explanation for differential growth rates is that, rather than greater transmissibility per se, B.1.1.7/VOC202012/01 may be more likely to lead to infection following any given exposure, consistent with enhanced ACE2 receptor binding associated with the N501Y mutation^3^. This hypothesis requires further investigation; whilst ACE2 gene expression increases with age^25^, effects could still be proportionate across the ages.

The main study strength is its design, being a large-scale community survey with a robust sampling frame across all ages and including asymptomatic infections, providing direct population-level estimates of positivity, in contrast to previous studies relying on routine symptomatic surveillance and necessary additional assumptions about parameter estimates based on non-random samples from other countries and time periods^6-8^. The main limitation is that not all SGTF will be B.1.1.7/VOC202012/01, even in the most recent period, as illustrated by varying Ct in SGTF by region (Figure 1B). Rather, we model SGTF in their entirety to estimate a “background” rate on which we can assess when B.1.1.7/VOC202012/01 might have arisen, as indicated by evidence for a change in trend using ISR. Enhanced whole genome sequencing for survey positives started mid-December, but was previously sparse. Preliminary data support B.1.1.7/VOC202012/01 comprising most SGTF in the survey from mid-November, and >88% of SGTF in the national symptomatic testing program are B.1.1.7/VOC202012/01 from this time^8^. However, conversely, some “non-SGTF” single N-gene or ORF1ab only positives (with high Ct^13^) could also be B.1.1.7/VOC202012/01. ISR does not identify “optimal” changepoints, and therefore does not correspond exactly to MRP, but this would be expected to lead to dilution bias. Analyses by participant do not account for within-household clustering; however, results were similar in household-level analyses (Supplementary Figure 4). Our analysis is based on regions, although there were some local differences in restrictions on hospitality and socialising within regions. However, mathematical models including only changes in behaviour/contact patterns poorly fitted observed data, suggesting this may have had less effect^6^.

In summary, direct representative population-level estimates of positivity across ages show that the new B.1.1.7/VOC202012/01 SARS-CoV-2 variant leads to higher infection rates overall, but is not particularly adapted to any specific age group. Although similar to other strains, the high percentage of infections without any evidence of symptoms, coupled with higher transmissibility, has made control extremely challenging without widespread whole community measures e.g. national lockdowns, school closures. Careful monitoring for ingress of B.1.1.7/VOC202012/01 through whole genome sequencing should be a priority in other countries. Continued surveillance for sudden increases in positivity rates which could herald its arrival, or that of other variants with enhanced transmissibility, remains essential^26^.

## Data Availability

De-identified study data are available for access by accredited researchers in the ONS Secure Research Service (SRS) for accredited research purposes under part 5, chapter 5 of the Digital Economy Act 2017. For further information about accreditation, contact or visit the SRS website.

## DATA AVAILABILITY

De-identified study data are available for access by accredited researchers in the ONS Secure Research Service (SRS) for <u>accredited research purposes</u> under part 5, chapter 5 of the Digital Economy Act 2017. For further information about accreditation, contact or visit the <u>SRS website.</u>

## CONTRIBUTIONS

The study was designed and planned by ASW, JF, JB, JN, IB, ID and KBP, and is being conducted by ASW, IB, RS and ER. This specific analysis was designed by ASW, KDV, OG and KBP. ASW, KDV, OG, EP, JJ and KBP contributed to the statistical analysis of the survey data. ASW drafted the manuscript and all authors contributed to interpretation of the data and results and revised the manuscript. All authors approved the final version of the manuscript.

## ACKNOWLEDGEMENTS

We are grateful for the support of all COVID-19 infection survey participants.

## Office for National Statistics

Sir Ian Diamond, Iain Bell, Emma Rourke, Ruth Studley, Alex Lambert, Tina Thomas.

**Office for National Statistics COVID Infection Survey Analysis and Operations teams**, in particular Daniel Ayoubkhani, Russell Black, Antonio Felton, Megan Crees, Joel Jones, Lina Lloyd, Esther Sutherland.

### University of Oxford, Nuffield Department of Medicine

Ann Sarah Walker, Derrick Crook, Philippa C Matthews, Tim Peto, Emma Pritchard, Nicole Stoesser, Karina-Doris Vihta, Alison Howarth, George Doherty, James Kavanagh, Kevin K Chau, Stephanie B Hatch, Daniel Ebner, Lucas Martins Ferreira, Thomas Christott, Brian D Marsden, Wanwisa Dejnirattisai, Juthathip Mongkolsapaya, Sarah Hoosdally, Richard Cornall, David I Stuart, Gavin Screaton.

### University of Oxford, Nuffield Department of Population Health

Koen Pouwels.

### University of Oxford, Big Data Institute

David W Eyre.

### University of Oxford, Radcliffe Department of Medicine

John Bell.

### Oxford University Hospitals NHS Foundation Trust

Stuart Cox, Kevin Paddon, Tim James.

### University of Manchester

Thomas House.

### Public Health England

John Newton, Julie Robotham, Paul Birrell.

### IQVIA

Helena Jordan, Tim Sheppard, Graham Athey, Dan Moody, Leigh Curry, Pamela Brereton.

### National Biocentre

Ian Jarvis, Anna Godsmark, George Morris, Bobby Mallick, Phil Eeles.

### Glasgow Lighthouse Laboratory

Jodie Hay, Harper VanSteenhouse.

