## Supplementary material for "Increased infections, but not viral burden, with a new SARS-CoV-2 variant"

**Supplementary Materials**

**Supplementary Methods**

As the cycle threshold (Ct) distribution was skewed, we assessed independent predictors using median (quantile) regression. We used 5 knot natural cubic splines (knots spaced 10%/25%/50%/75%/90% from the minimum to the maximum) to assess non-linearity in the effect of calendar time. Analyses were conducted in Stata 16.1.

We analysed the percentage of the private-residential population testing positive for SARS-CoV-2 from nose and throat swabs over time using Bayesian dynamic multi-level regression and post-stratification (MRP)[1,2] to correct for any residual non-representativeness in terms of age, sex and region. Several empirical and simulation studies have found MRP to be superior to classical survey weighted and unweighted approaches, including when using small sample sizes at national and regional levels[1-3]. Partial pooling through the use of random effects in the multilevel model ensures stable estimates can be obtained for subnational levels from relatively small samples that would be problematic using more traditional survey-weighting approaches. Multilevel generalised additive regression was used to model the swab test result as a function of age, sex, time and region. Separate models were fitted for S-gene target failure (SGTF) positives (positive on only ORF1ab and the N-gene) vs other results (negative or non-SGTF positive) and non-SGTF positives vs other results (negative or SGTF). Time, measured in days since 28 September 2020, was modelled using thin-plate splines and allowed to vary by region. We set k, the number of basis functions, to 10 to control the smoothness of the fitted function[4]. We used a normal prior with location set to 4 for the standard deviation of the smooth, as previously[5]. Subsequently, we post-stratified the resulting positivity estimates for each demographic-geographic respondent type by the percentage of each type in the overall population and in each region. We did not post-stratify for other factors (e.g. ethnicity) because reliable estimates in the target population were not available. We fitted separate models to the nine English regions and each of the three devolved administrations. Because there were very few missing values (≤1%) in age and sex, we restricted all analyses to observations with non-missing data. A complementary log-log link was used due to the ability to interpret regression coefficients as arising from an infection process with varying levels of exposure[6]. MRP models with random effects for individual participant and/or household nested within region did not converge. Therefore MRP models were run with only a random intercept for region, without a random intercept for participant and/or household. Models with only one participant sampled from each household have given similar results in previous analyses with somewhat wider 95% credible intervals mainly due to the smaller sample size[5]. Analyses were performed using the rstanarm package in R version 3.6.1.19.

To estimate current growth rates in SGTF and non-SGTF positives, we used the iterative Sequential Regression (ISR) algorithm[7,8] to estimate changepoints in unweighted positivity estimates over calendar time. Growth rates (-ln(2)/ln(rate ratio per day)) can then be estimated directly from the most recent trend. In summary, ISR looks at the data iteratively, and compares models with one trend with those with 2 trends; if based on some criterion, the one with 2 trends is a better fit, then the "changepoint" is fixed and the process is repeated (i.e. more data is added and new models with this changepoint, but also other potential changepoints after this initial one are fitted). This method enables an unknown number of multiple changes in trend to be estimated efficiently, in contrast to traditional grid search algorithms which require the number of changes in trend to be fixed, and, in addition, also require every possible combination of changepoints to be modelled, making them very computationally intensive even for a small number of changepoints. We considered the binary outcome positive of specific type (1) vs negative or other positive (0) using a log link and poisson regression. The ISR algorithm first modelled the outcome using swab results from the 1st of September to the 22nd of September, and compared a model with one trend over calendar time in the outcome to a model allowing this trend to change on the 11th of September (keeping a minimum of 10 days from the last changepoint to the last data included in the model). If the model with two trajectories was not a better fit (determined by a Akaike Information Criterion being lower by at least 6.635 for non-SGTF positives as the outcome and 3.84 for SGTF positives (the critical value corresponding to a significance level of 0.01 or 0.05 with one degree of freedom, respectively, given the very different positivity rates)), then an additional 1 day's observations (to 23rd of September) were included. The model with one trend was then compared to models with 2 trajectories with changepoints on the 11th of September or the 12th of September (keeping a minimum of 10 days of data again to the last data included in the model), again considering whether any model with a change in trend substantially improved model fit. Any changepoint that improved model fit was fixed, and then an additional 20 days of data included (since only changepoints at least 10 days from both the previous changepoint and the last data included in the model were considered). This process was iterated up to the end of the data. Outputs are rate of change per day in the most recent period since the last change in trend was detected (at the last "changepoint") and the current positivity rate (percentage) for each type. Analyses were performed in R version 3.6.1.19. The average difference between log growth rates for SGTF vs non-SGTF positives were estimated using random-effects meta-analysis, assuming independence (i.e. taking the variance as the sum of the variances of the individual growth rates, since there is no straightforward method of estimating correlations between trends in different outcomes that change at different timepoints).

Analyses of growth rates by age divided the population into those up to and including 15/16 years (up to and including high school age) and older. School years rather than absolute years were used because behaviour is more likely to reflect place of education than numerical age for those 15-17 years.

The impact of age was further explored by analysing positivity rates over time by age as a continuous factor. Generalised additive models with a complementary loglog link and a tensor product thin plate splines were used to model, for each region separately, an interaction between continuous time (measured in days) and continuous age (measured in years). Separate models were used to estimate SGTF and non-SGTF positivity over time and by age. For thin-plate splines, *k* – the choice of the basis dimension - has to be set large enough in order to ensure the true variation of the true underlying function is accommodated, with the caveat that too large k-values can be computationally infeasible. The degree of penalisation to control the smoothness and avoid overfitting was controlled automatically by optimising the Un-Biased Risk Estimator (UBRE)[4,9]. Here we set k-values to 32 (for time, in total 126 days from 28 September) and 26 (for age, range 2-80).

**Supplementary Figure 1 Percentage of positives and Ct values over time, Devolved Administrations**

A Percentage of positives that were SGTF

(ORF1ab+N positive only)

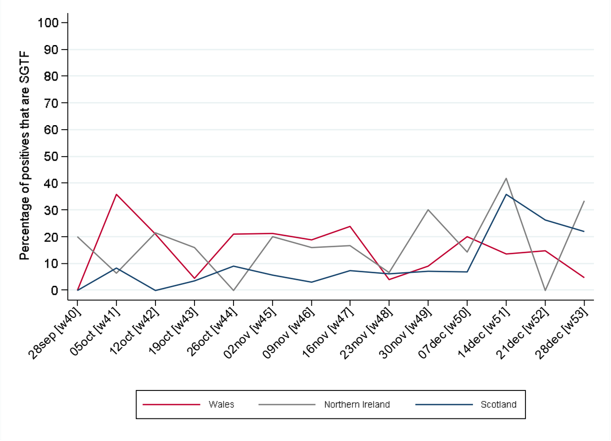


B Median Ct values in SGTF

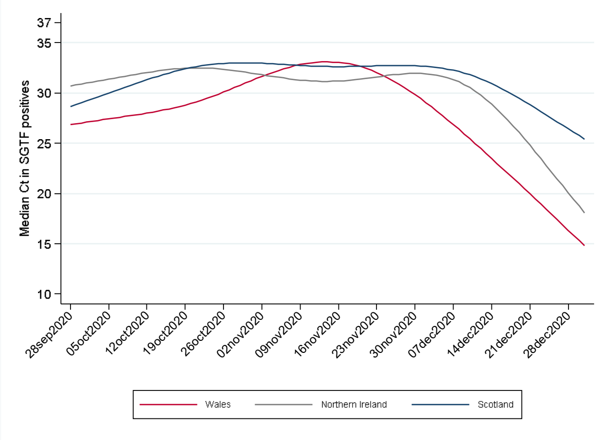


C Percentage of positives that were triple-gene

positive

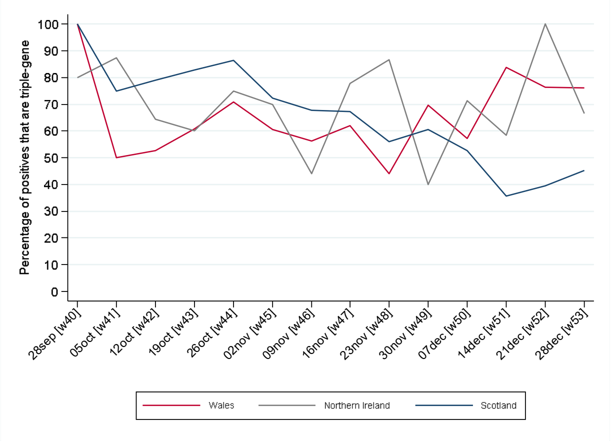


D Median Ct values in triple-gene positives

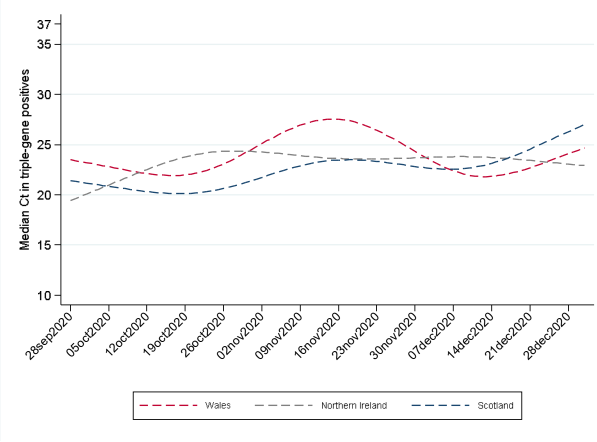

**Supplementary Figure 2 Percentage of positives with Ct<30 that were SGTF (ORF1ab+N positive) (A, B) or triple gene positive (C, D) in English regions (A, C) and Devolved Administrations (B, D)**

A English regions, SGTF (ORF1ab+N positive), Ct<30

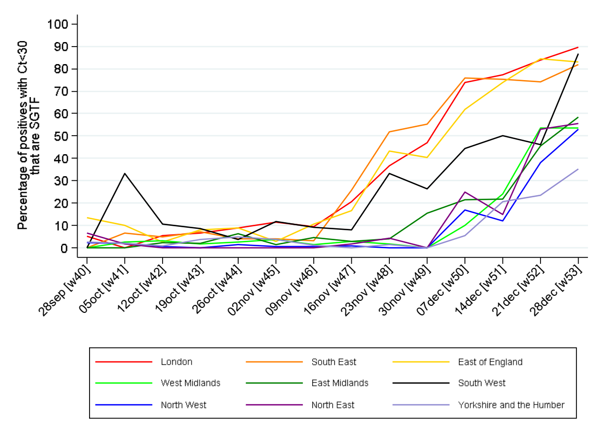


B Devolved Administrations, SGTF (ORF1ab+N positive), Ct<30

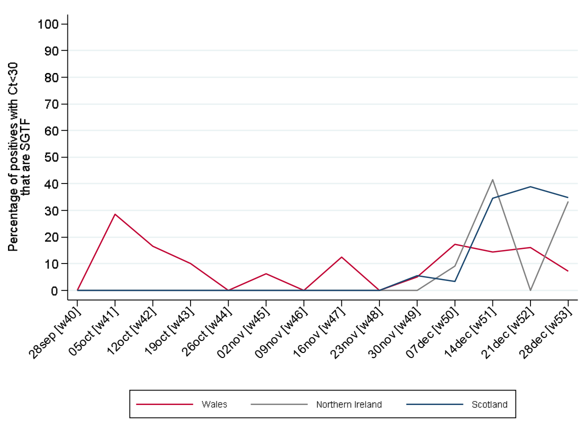


C English regions, triple-gene positives, Ct<30

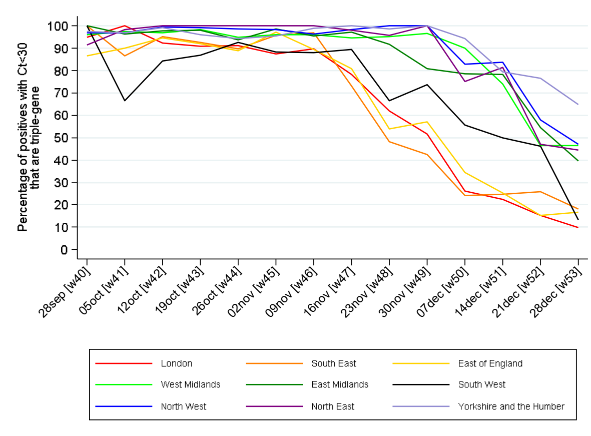


D Devolved Administrations, triple-gene positives, Ct<30

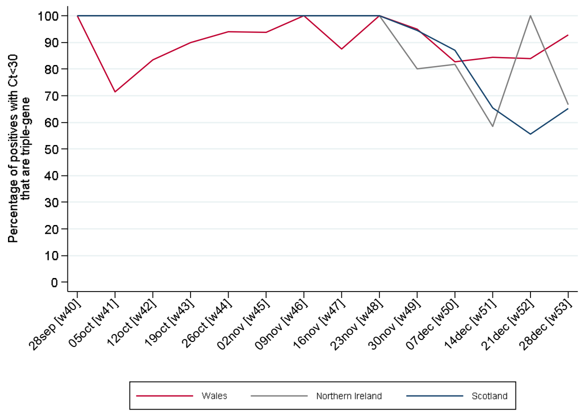

**Supplementary Figure 3 Percentage of the population positive with SGTF (ORF1ab+N positive, compatible with B.1.1.7/VOC202012/01) and non-SGTF – log scale**

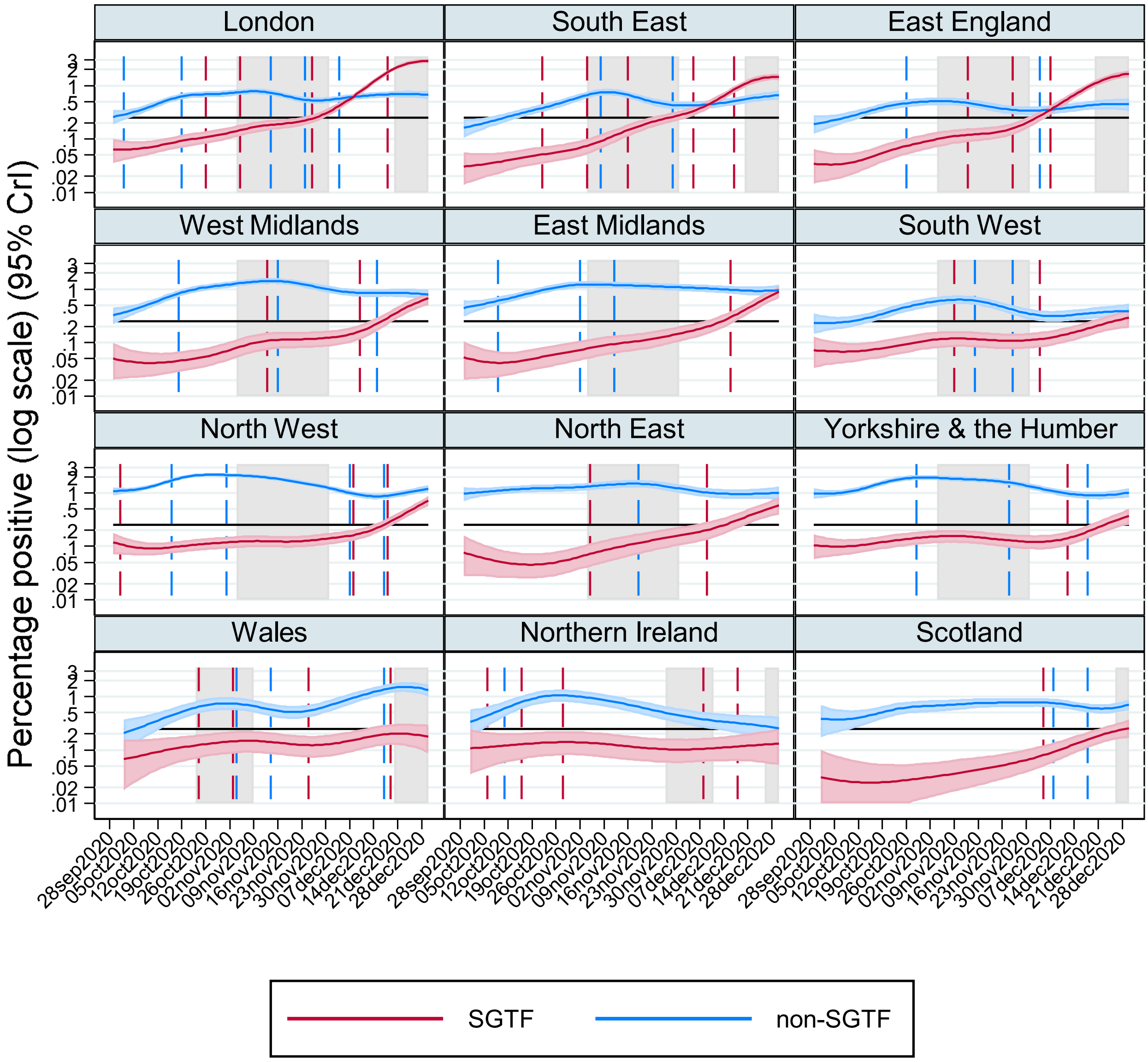


Note: truncating lower 95% CrI at 0.01. Gray shading shows national restrictions/stay at home orders for the majority of the region. Black horizontal line at 0.25%. Dashed lines show changes in trend from ISR algorithm fitted from 1 September (no dashed line means no change in trend with p<0.01 (non-SGTF) or p<0.05 (SGTF) detected). See **Figure 2** for probabilities on the absolute scale.

**Supplementary Figure 4 Percentage of households positive with SGTF (ORF1ab+N positive, compatible with B.1.1.7/VOC202012/01) and non-SGTF – absolute (A) and log (B) scale**

**(A)**

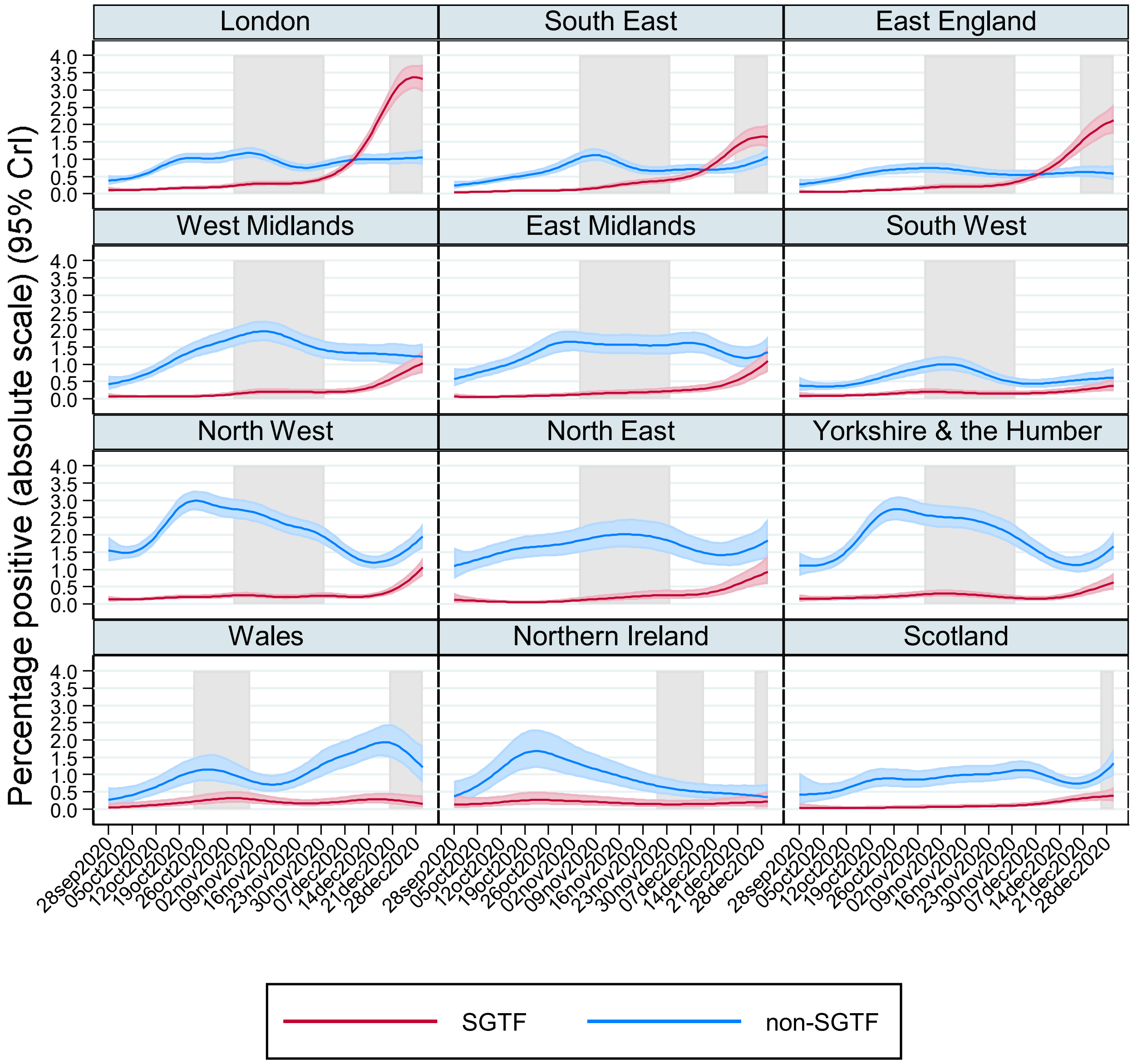


Note: gray shading shows national restrictions/stay at home for the majority of the region.

**(B)**

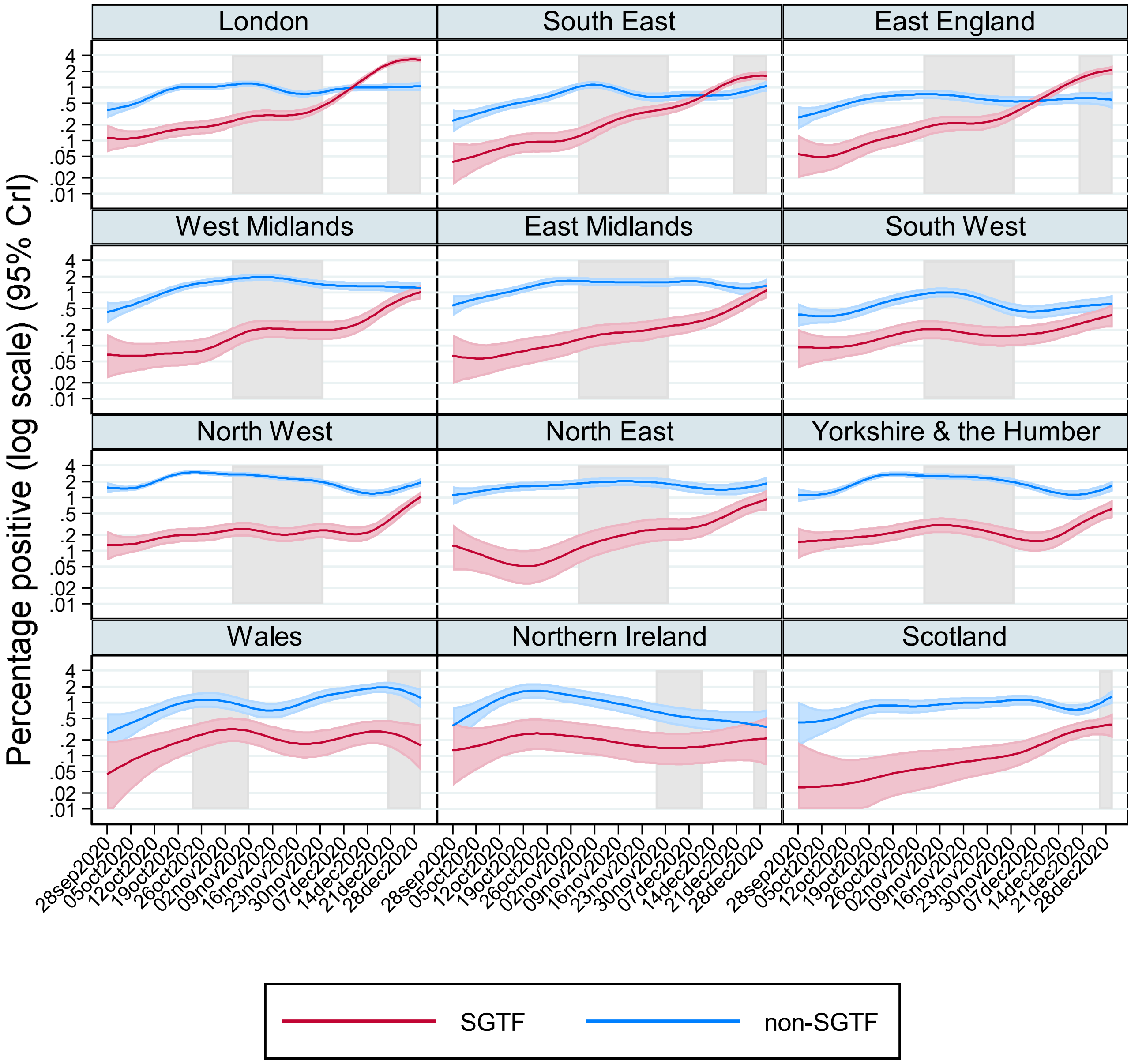


Note: truncating lower 95% credible interval at 0.01. Gray shading shows national restrictions/stay at home for the majority of the region.

**Supplementary Figure 5 Percentage of the population non-SGTF positive according to self-reported symptoms at the test**

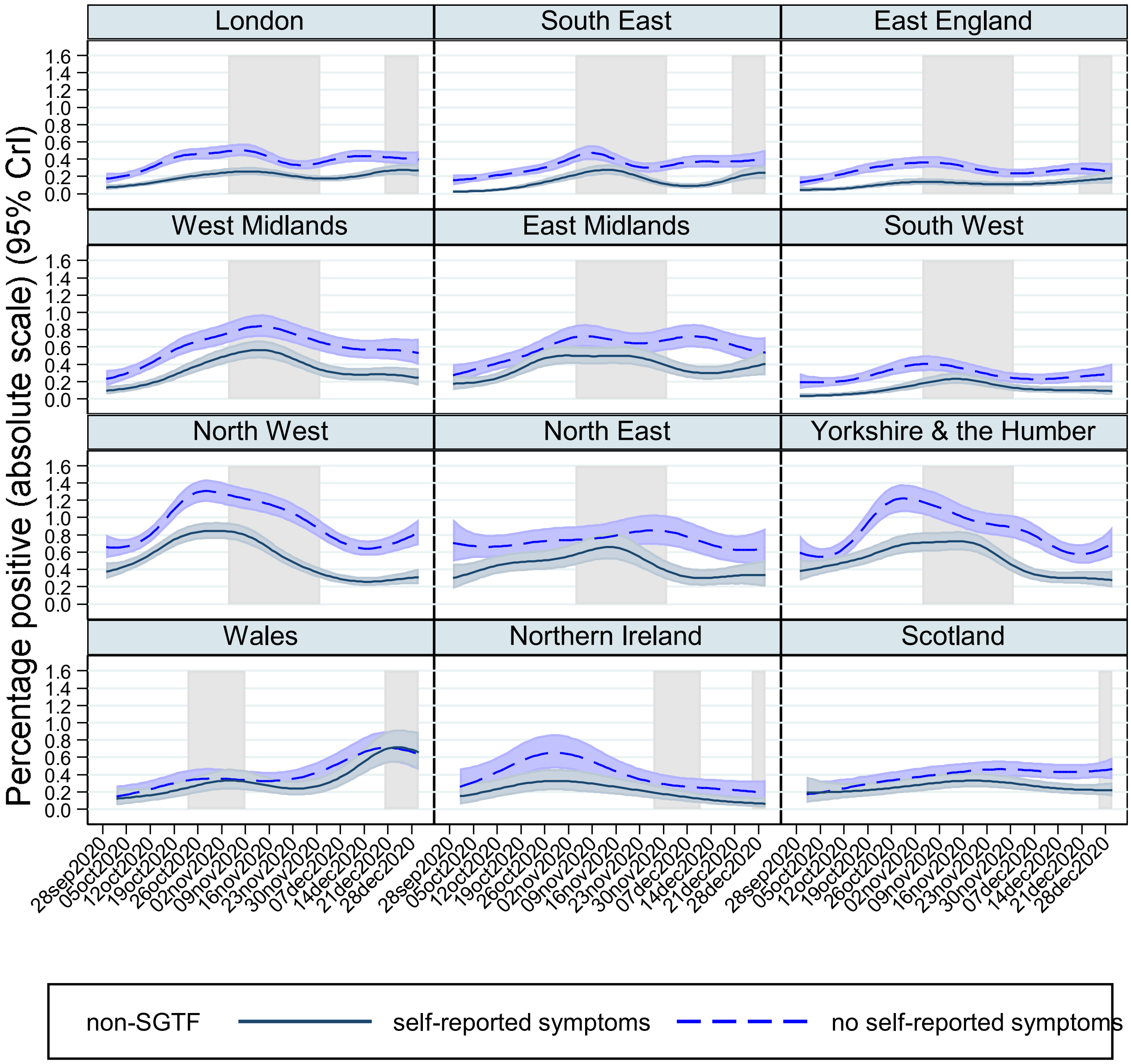


Note: gray shading shows national restrictions/stay at home for the majority of the region.

**Supplementary Figure 6 Percentage of the population SGTF (A) and non-SGTF (B) positive with Ct<30 according to self-reported symptoms at the test**

**(A)**

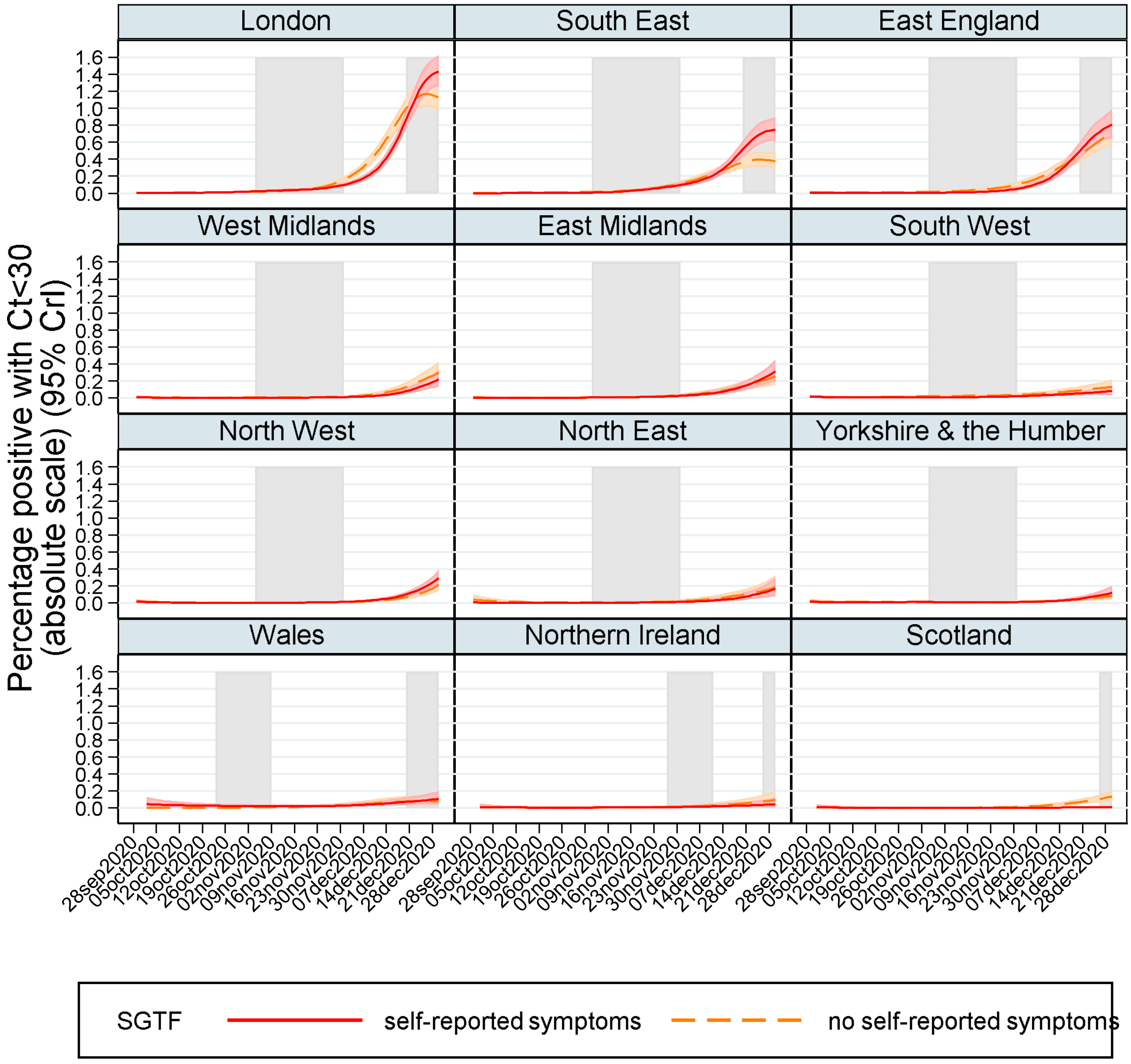


Note: gray shading shows national restrictions/stay at home for the majority of the region.

**(B)**

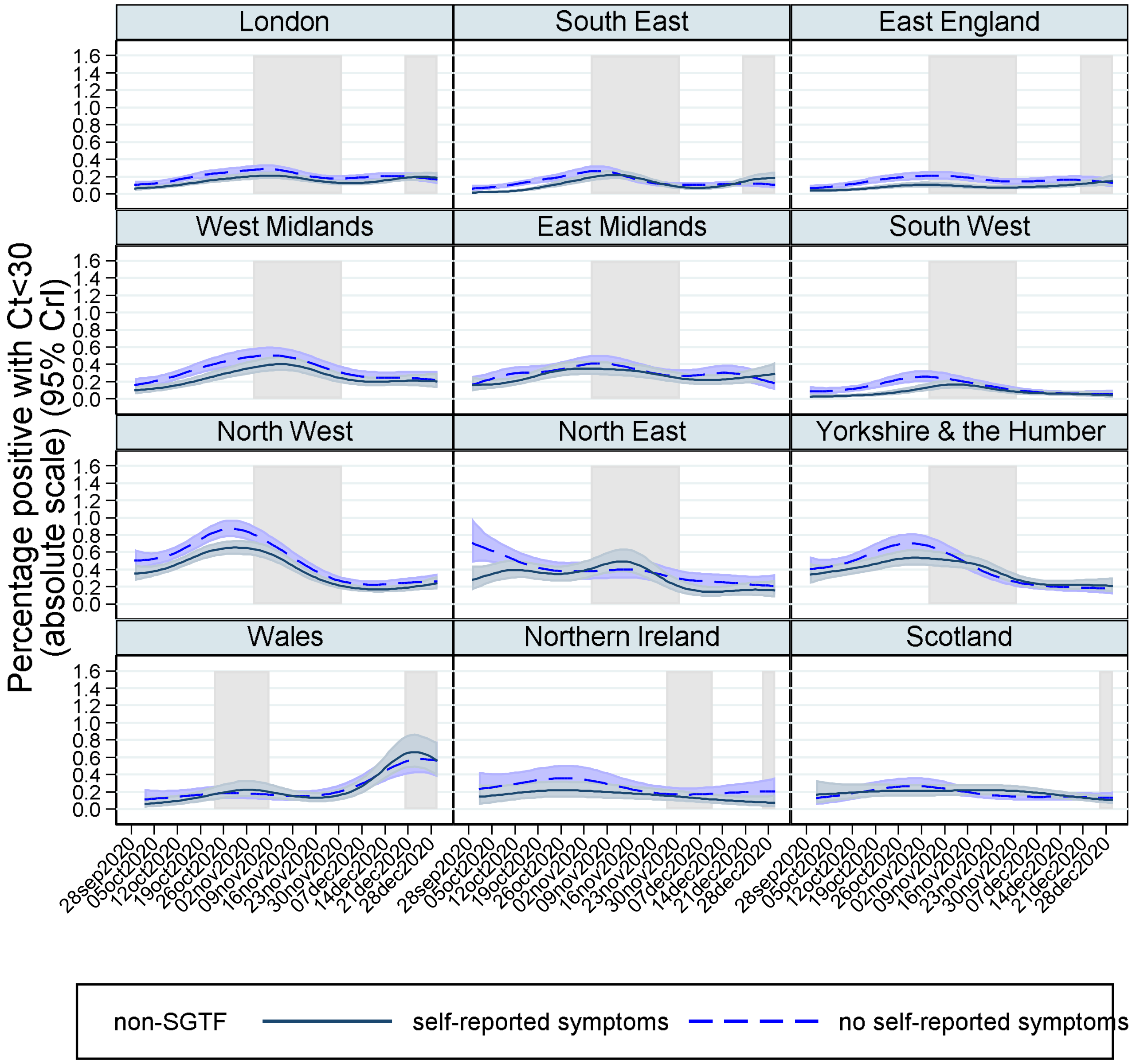


Note: gray shading shows national restrictions/stay at home for the majority of the region.

**Supplementary Figure 7 Rate of change in growth by absolute percentage positive**

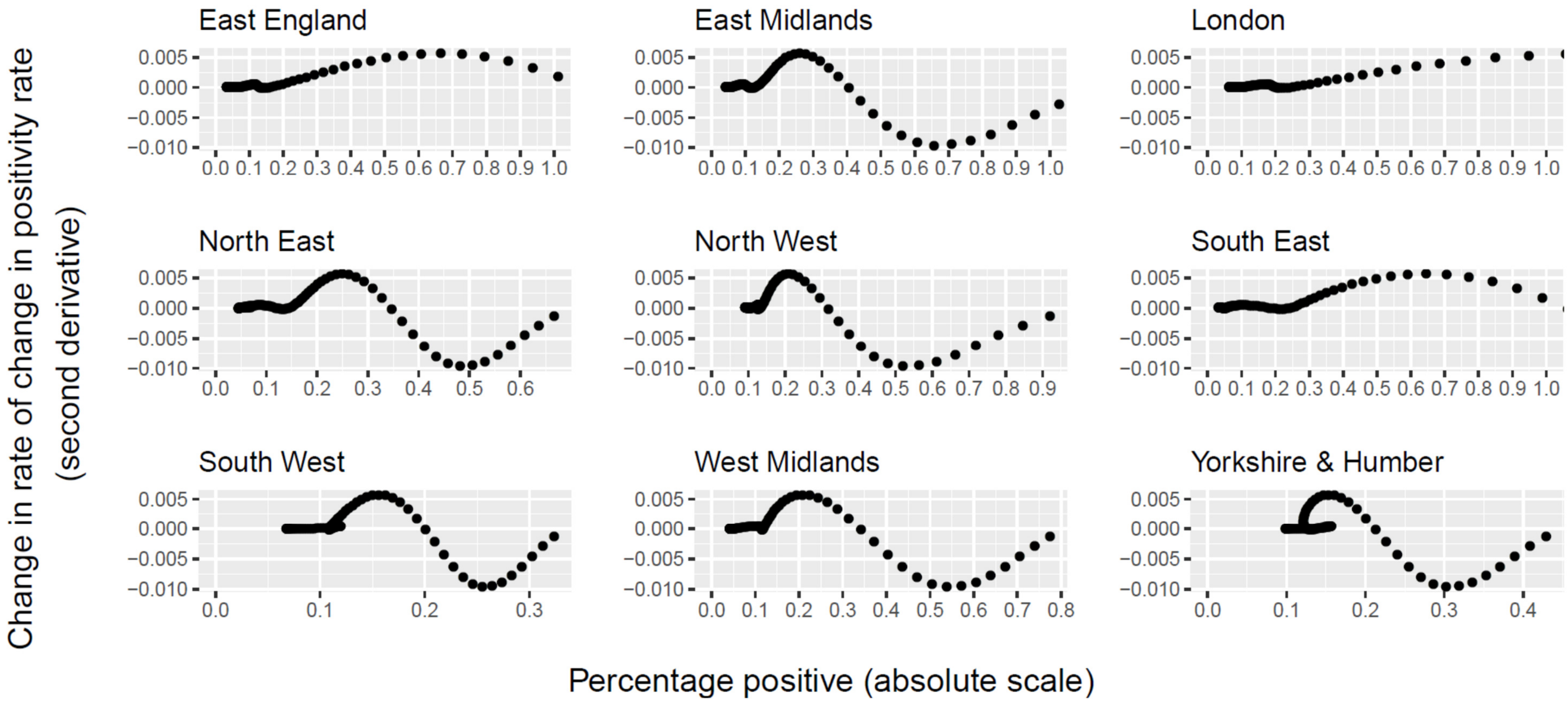


Note: showing the second derivative of the positivity rate estimated from MRP (**Figure 2**) by the estimated positivity rate. Higher values mean growth is accelerating faster, so the maximum provides some indication of when SGTF was increasing fastest.

**Supplementary Figure 8 Growth rates in SGTF vs non-SGTF in the most recent epoch (A) and the preceding epoch (B)**

**(A) most recent epoch**

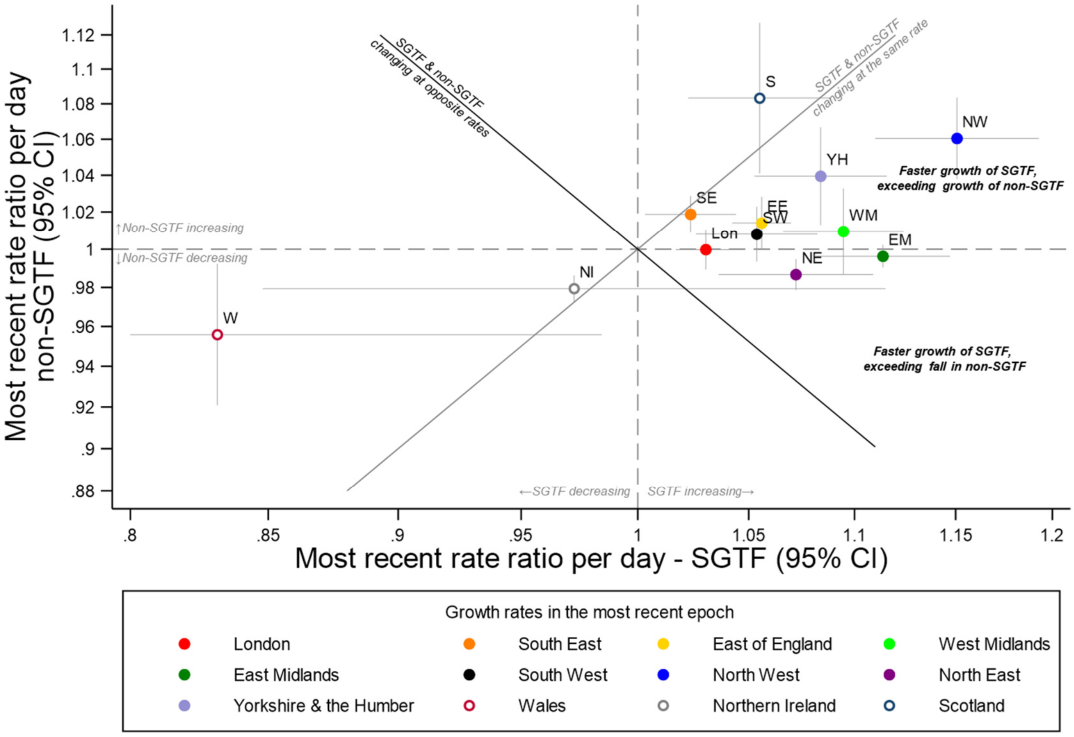


**(B) preceding epoch**

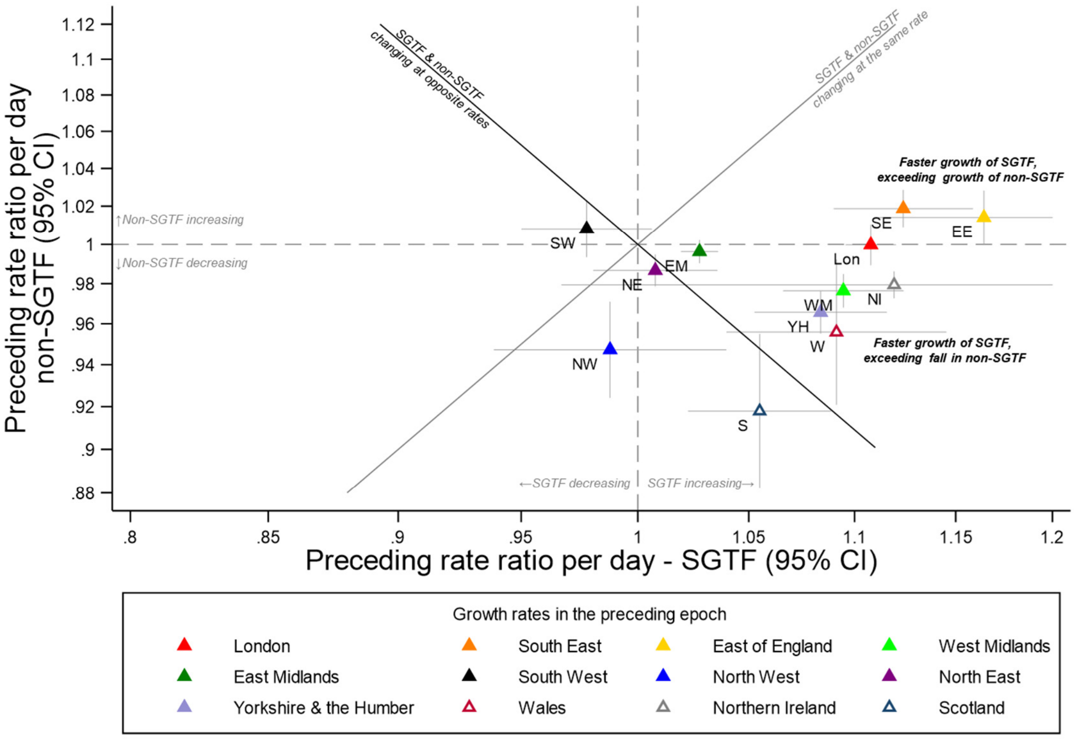

Note: panels (A) (B) shows growth rates (rate ratio (RR) per day) of SGTF (x-axis) and non-SGTF (y-axis) positives within the same region in epochs defined by changepoints (change in trend) identified by ISR and shown in **Figure 2** and **Supplementary Table 1**. 95% CI are truncated at 0.8 and 1.2. RR>1 mean positivity rates are increasing, <1 that they are decreasing. Points to the right of the gray diagonal line are periods of time where, within one region, SGTF positives are increasing faster than non-SGTF positives; and points on/around the gray line where SGTF and non-positives are changing at similar rates within a region. The black diagonal line indicates opposite growth rates, that is SGTF are increasing at the same rate non-SGTF are decreasing or vice versa within a region, consistent with replacement. Points above the black line are consistent with addition. Points from both panels are combined in **Figure 4A**.

**Supplementary Figure 9 Growth rates (X-axis) and estimated percentage positive (y-axis) for SGTF (A) and non-SGTF (B) in January 2021**

**(A) SGTF**

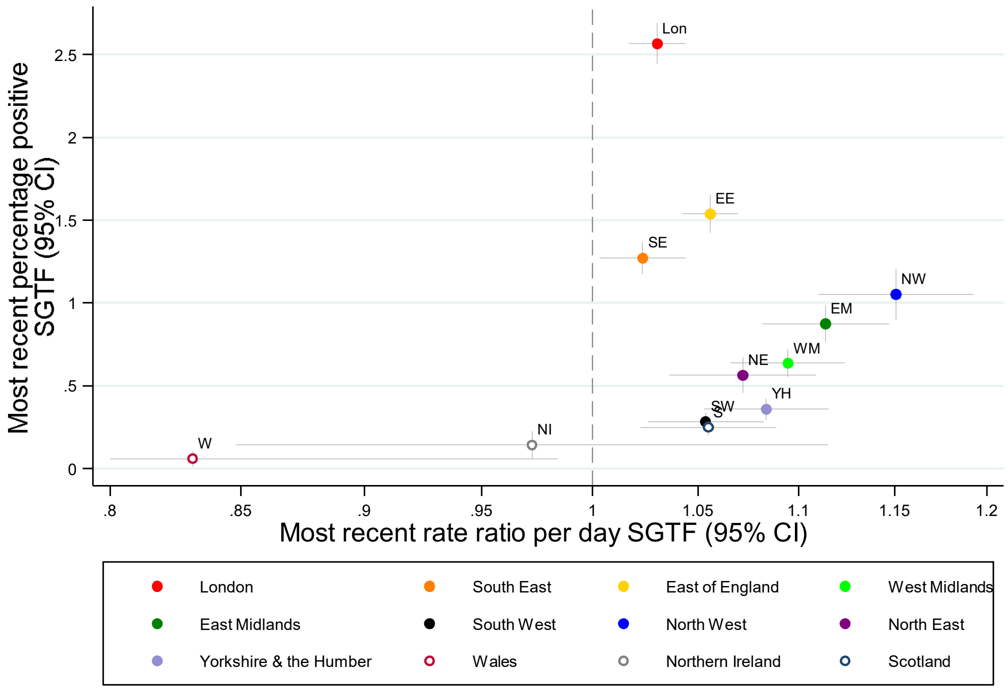


Note: truncating rate ratio 95% CI at 0.8

**(B) non-SGTF**

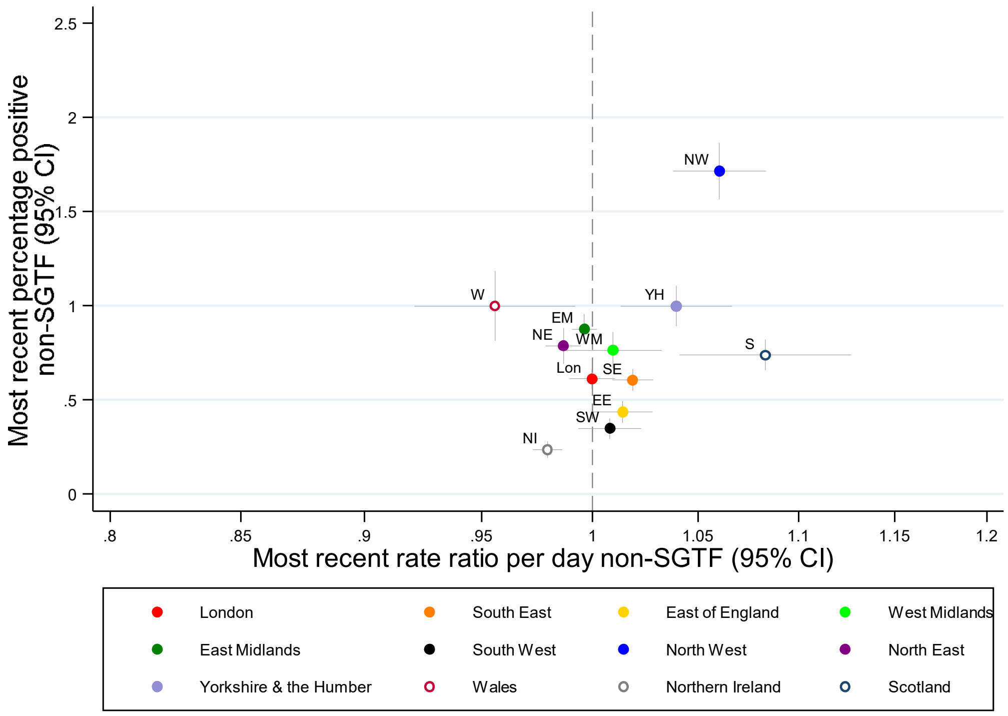

Note: panels show estimates of the positivity rate with 95% CI on the y-axis and the current rate ratio (RR) (growth rate) (95% CI on the x-axis). RR>1 mean positivity rates are increasing, <1 that they are decreasing. Points in the upper right quadrant are regions where current rates of this type of positive are high and increasing.

**Supplementary Figure 10 Growth rates of SGTF and non-SGTF positives with Ct<30 in two most recent epochs defined by ISR**

**(A) growth rates for positives with Ct<30**

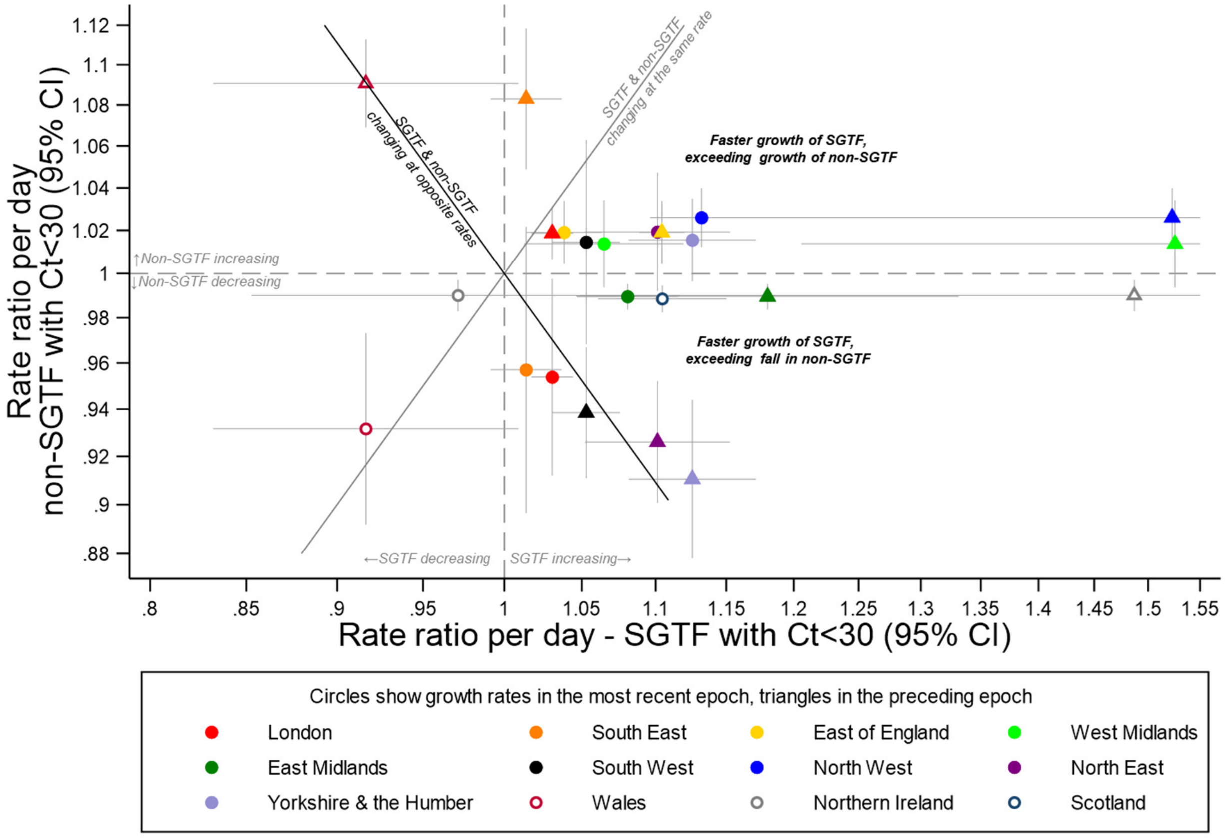

**(B) difference in growth rates for positives with Ct<30**

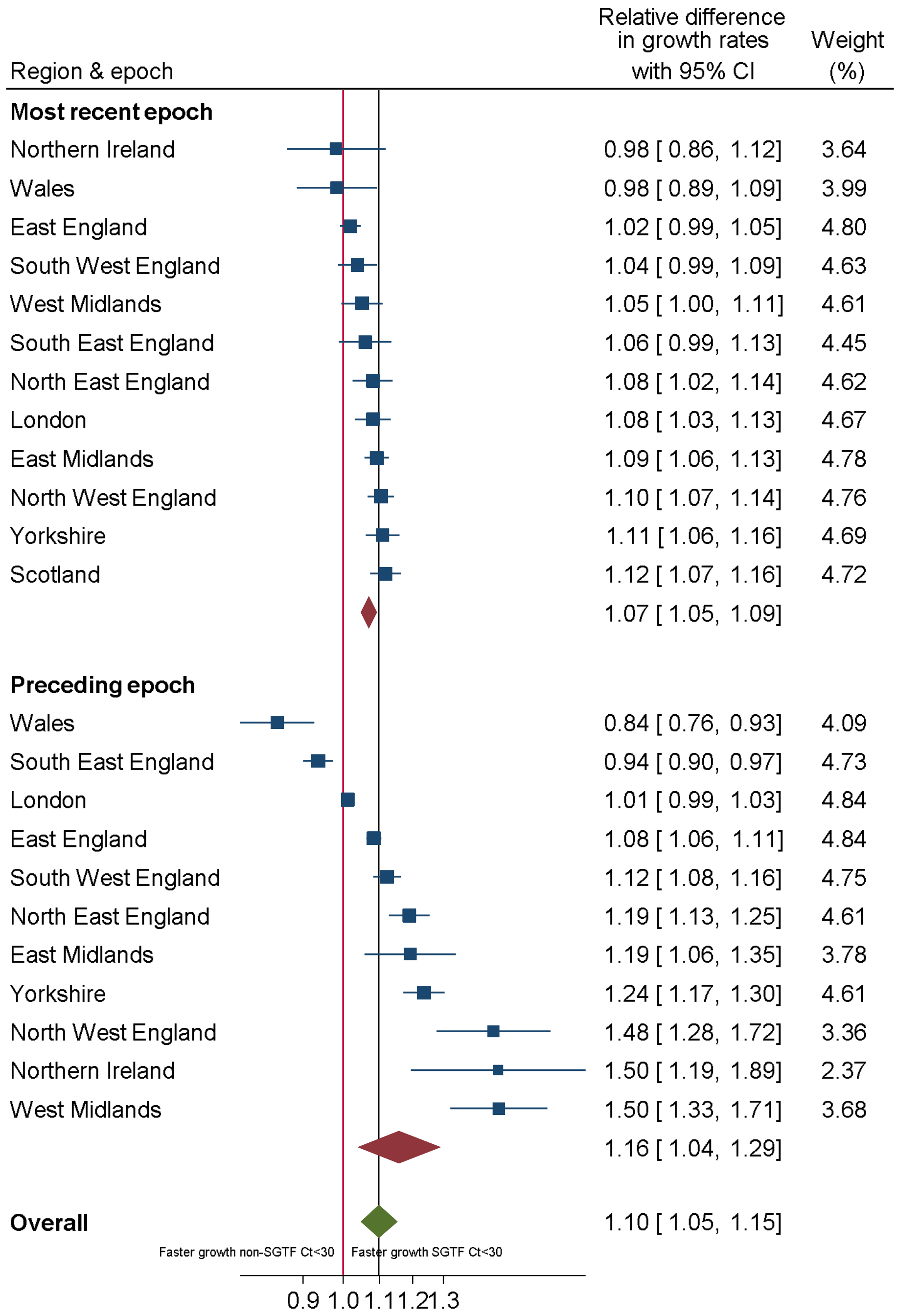


Note: panel (A) shows growth rates (rate ratio (RR) per day) of SGTF (x-axis) and non-SGTF (y-axis) positives **with Ct<30** within the same region in epochs defined by changepoints (change in trend) identified by ISR. 95% CI are truncated at 0.8 and 1.55. RR>1 mean positivity rates are increasing, <1 that they are decreasing. Points to the right of the gray diagonal line are periods of time where, within one region, SGTF positives are increasing faster than non-SGTF positives; and points on/around the gray line where SGTF and non-positives are changing at similar rates within a region. The black diagonal line indicates opposite growth rates, that is SGTF are increasing at the same rate non-SGTF are decreasing or vice versa within a region, consistent with replacement. Points above the black line are consistent with addition. Panel B shows difference between growth rates in SGTF and non-SGTF from panel (A), combined using random effects meta-analysis.

**Supplementary Figure 11 Growth rate in SGTF vs non-SGTF in those up to high school age versus older in the two most recent epochs (A), the most recent epoch (B) and the preceding epoch (C), and differences in growth rates in those aged up to high school (D) and older (E)**

**(A) two most recent epochs**

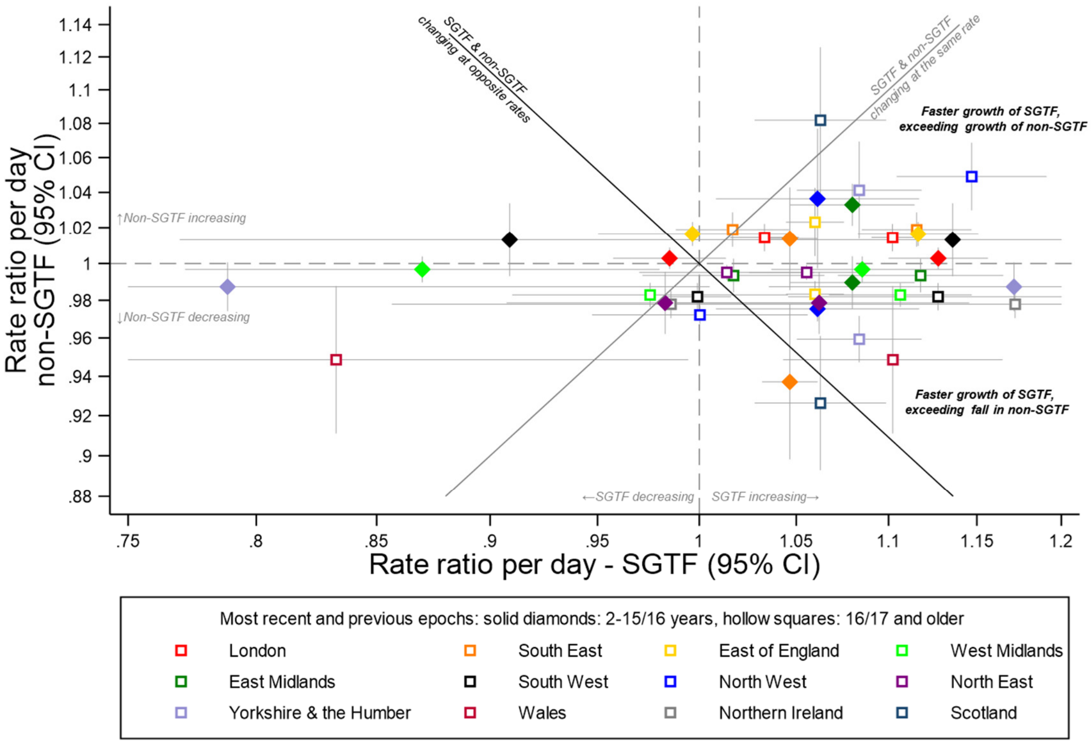


**(B) most recent epoch**

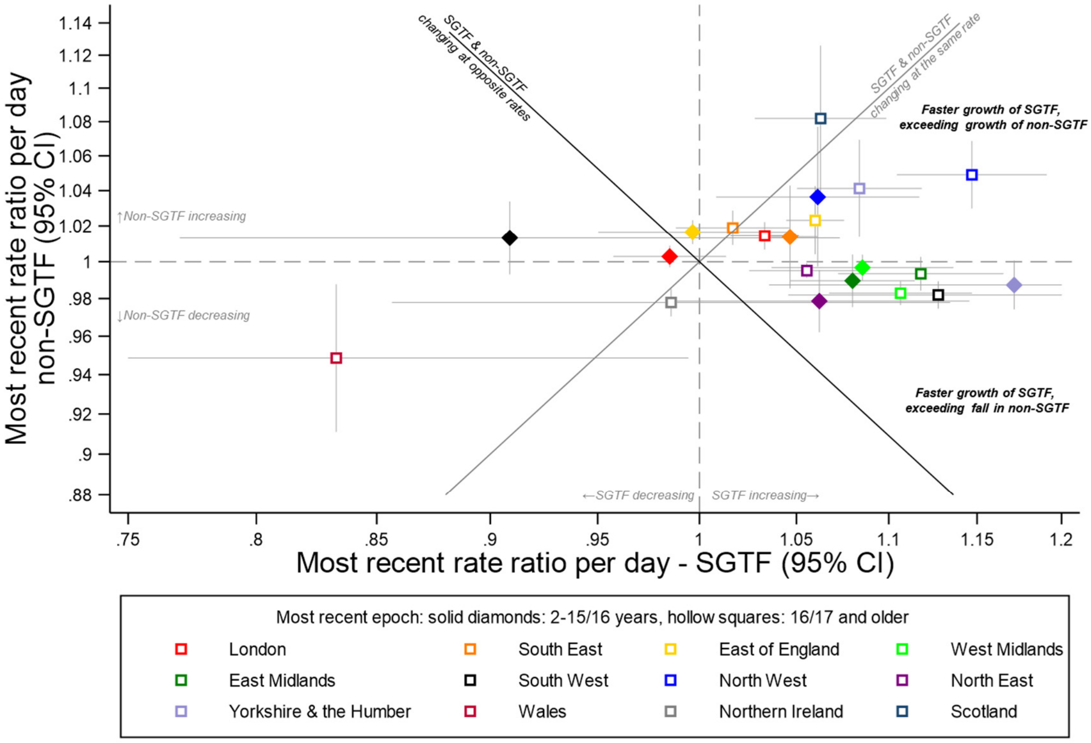

**(C) preceding epoch**

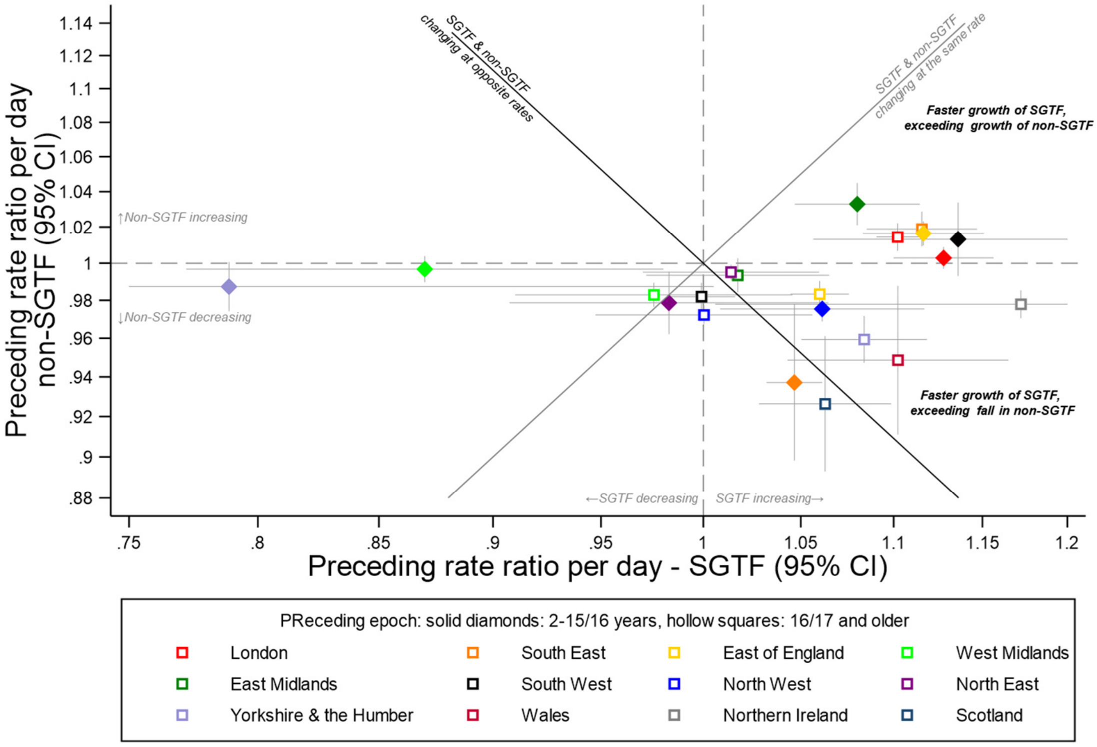

**(D) Difference in growth rates SGTF vs non-SGTF in those aged up to high school age**

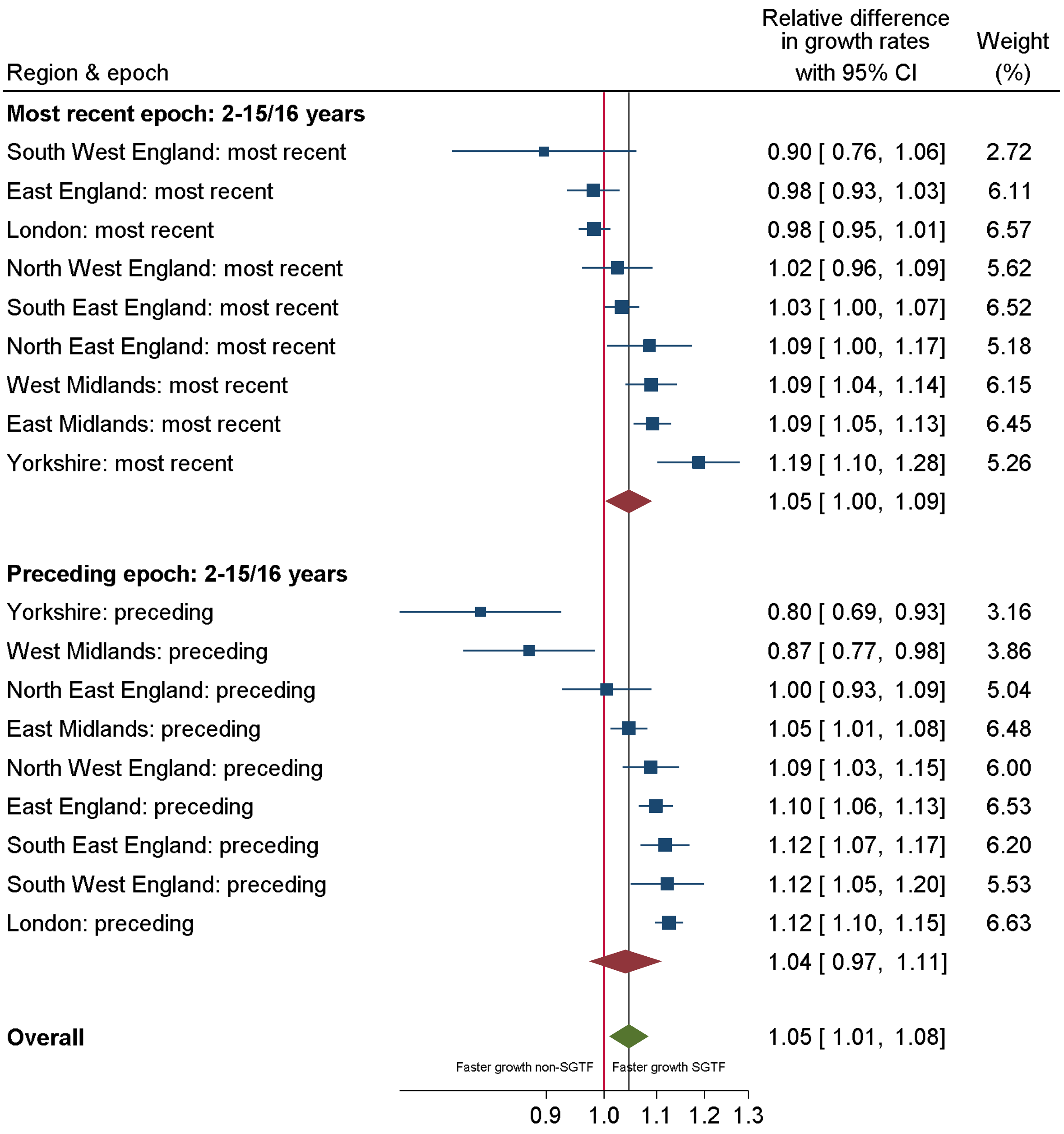

**(E) Difference in growth rates SGTF vs non-SGTF in those aged over high school age**

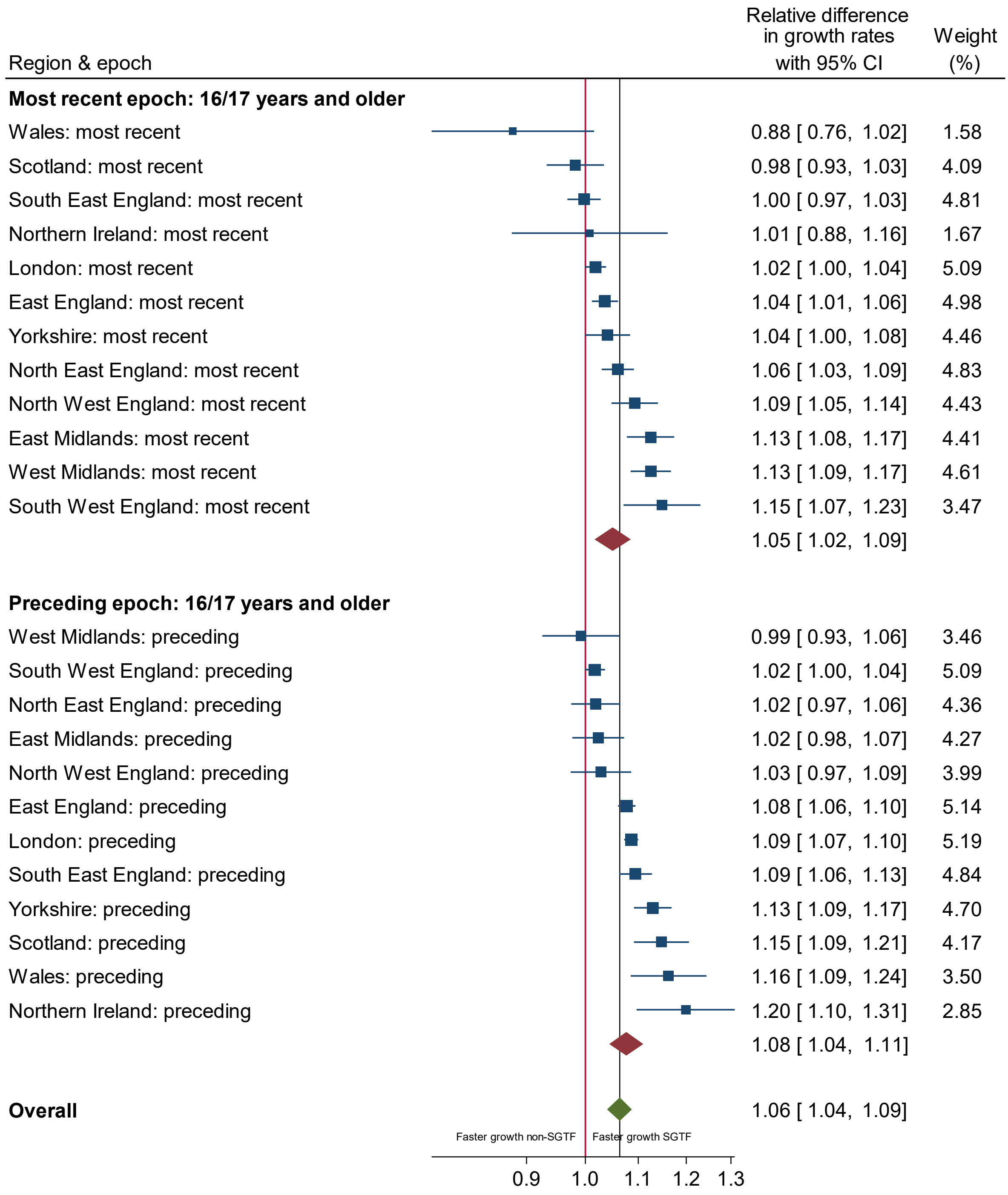

Note: panels (A) (B) (C) shows growth rates (rate ratio (RR) per day) of SGTF (x-axis) and non-SGTF (y-axis) positives within the same region in epochs defined by changepoints (change in trend) identified by ISR and shown in **Supplementary Table 2**. 95% CI are truncated at 0.75 and 1.2. RR>1 mean positivity rates are increasing, <1 that they are decreasing. Points to the right of the gray diagonal line are periods of time where, within one region, SGTF positives are increasing faster than non-SGTF positives; and points on/around the gray line where SGTF and non-positives are changing at similar rates within a region. The black diagonal line indicates opposite growth rates, that is SGTF are increasing at the same rate non-SGTF are decreasing or vice versa within a region, consistent with replacement. Points above the black line are consistent with addition. Panels D and E shows difference between growth rates in SGTF and non-SGTF from panels A, B, C, combined using random effects meta-analysis.

**Supplementary Figure 12 Impact of age on SGTF and non-SGTF growth rate**

**(A) non-SGTF growth rates in younger and older individuals**

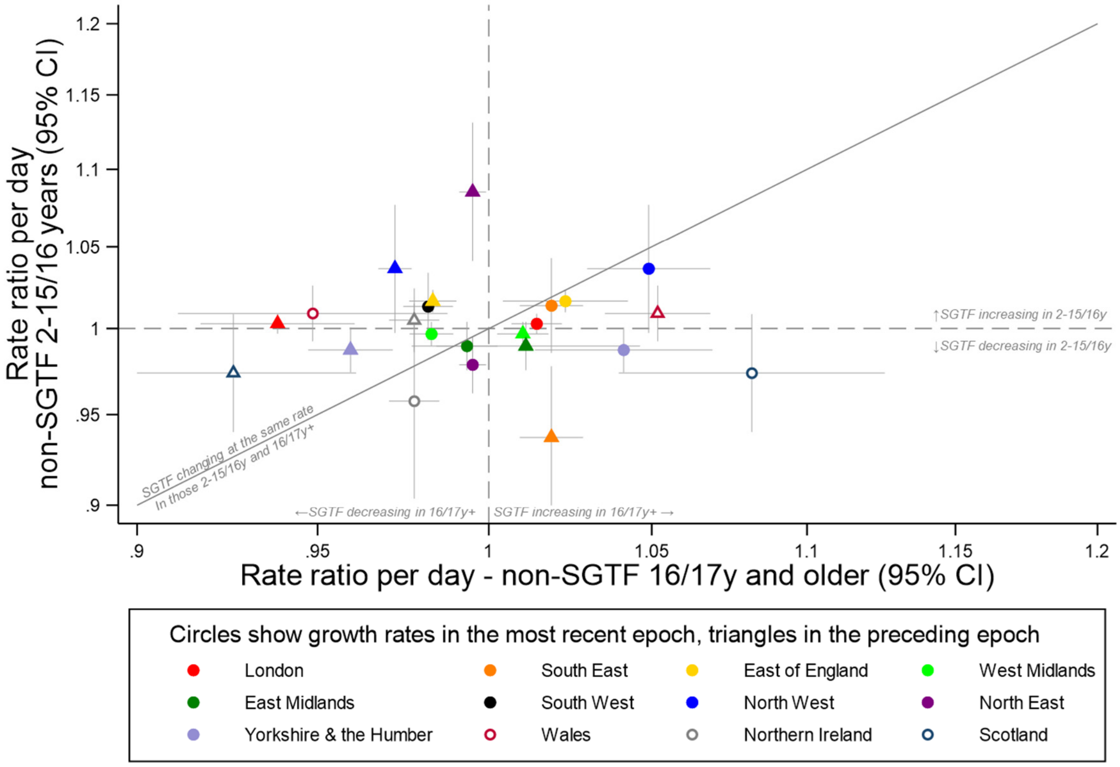

**(B) Difference in growth rates for SGTF between younger and older individuals**

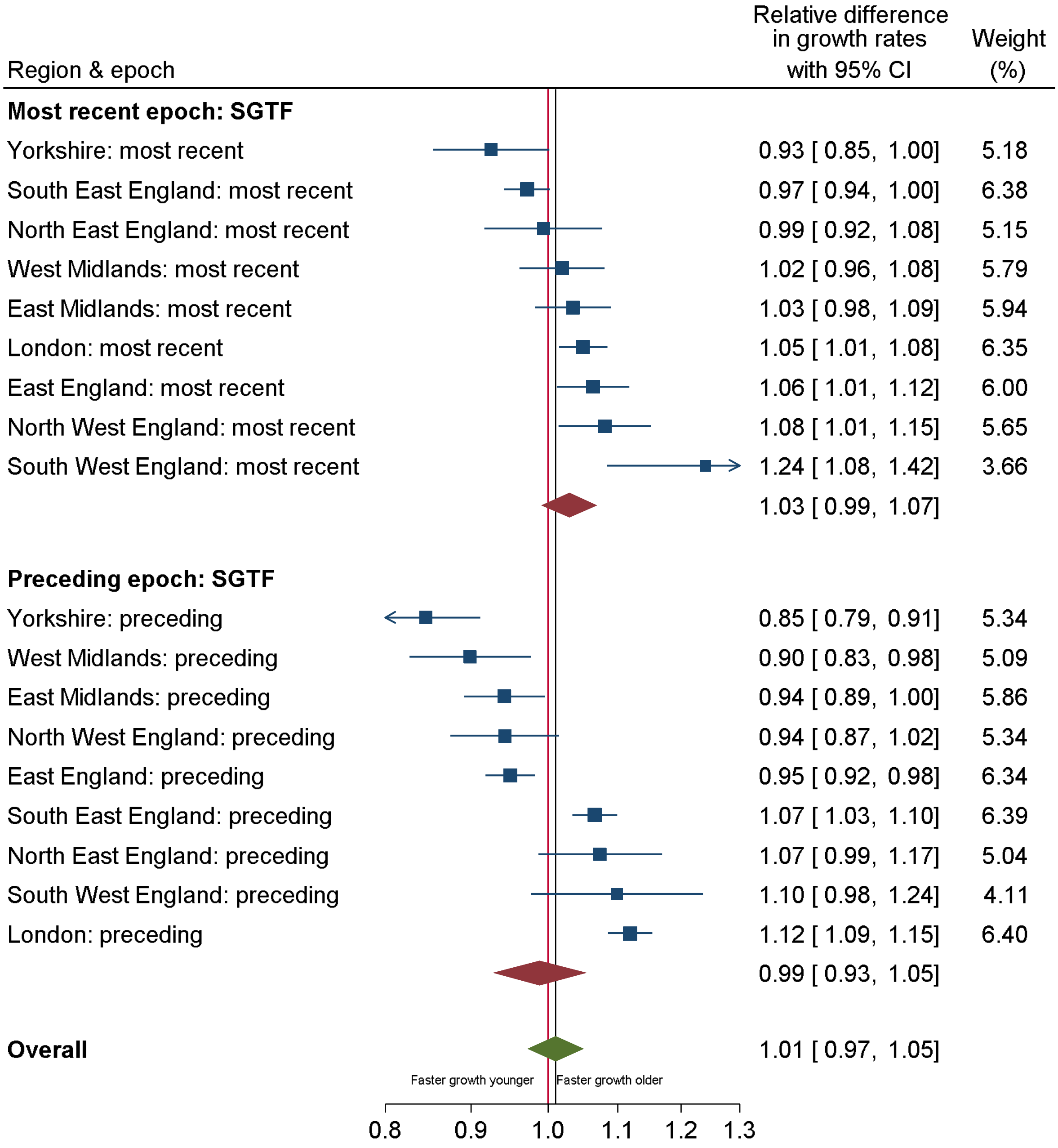

**(C) Difference in growth rates for non-SGTF between younger and older individuals**

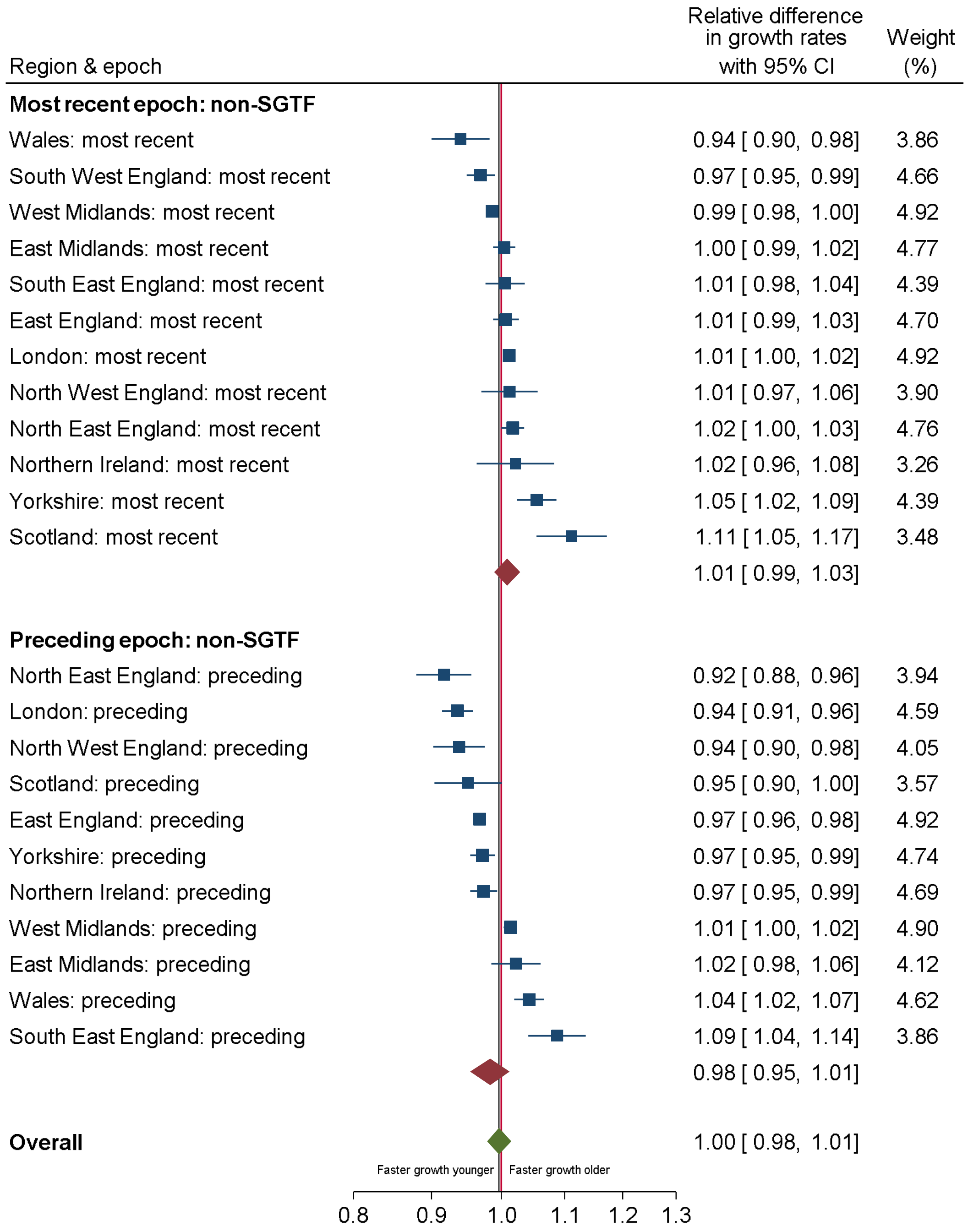


Note: panel A shows growth rates (rate ratio (RR) per day) of non-SGTF in 16/17y and older (x-axis) and of SGTF in those aged 2 to 15/16 years (y-axis) positives within the same region in epochs defined by changepoints (change in trend) identified by ISR and shown in **Supplementary Table 2**. 95% CI are truncated at 0.85 and 1.2. RR>1 mean positivity rates are increasing, <1 that they are decreasing. Points to the right of the gray diagonal line are periods of time where, within one region, non-SGTF positives are increasing faster in older than younger individuals; and points on/around the gray line where non-SGTF positives are changing at similar rates within a region in both younger and older individuals. Panels B and C show differences between growth rates in SGTF (B) (corresponding to **Figure 5A**) and non-SGTF (C) (corresponding to panel A) growth rates in younger vs older individuals combined using random effects meta-analysis.

**Supplementary Figure 13 Percentage of positives that were SGTF by age and region 21 December 2020-2 January 2021**

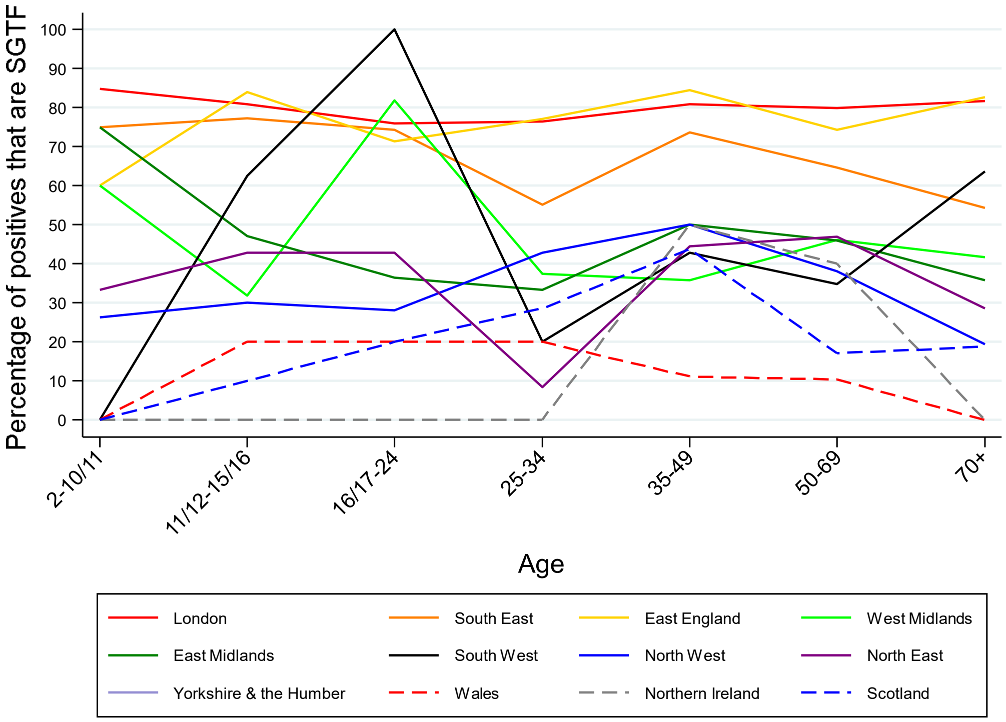


Note: raw data in **Supplementary Table 3**.

**Supplementary Figure 14 Percentage of positives that were SGTF by age and region 21 December 2020-2 January 2021**

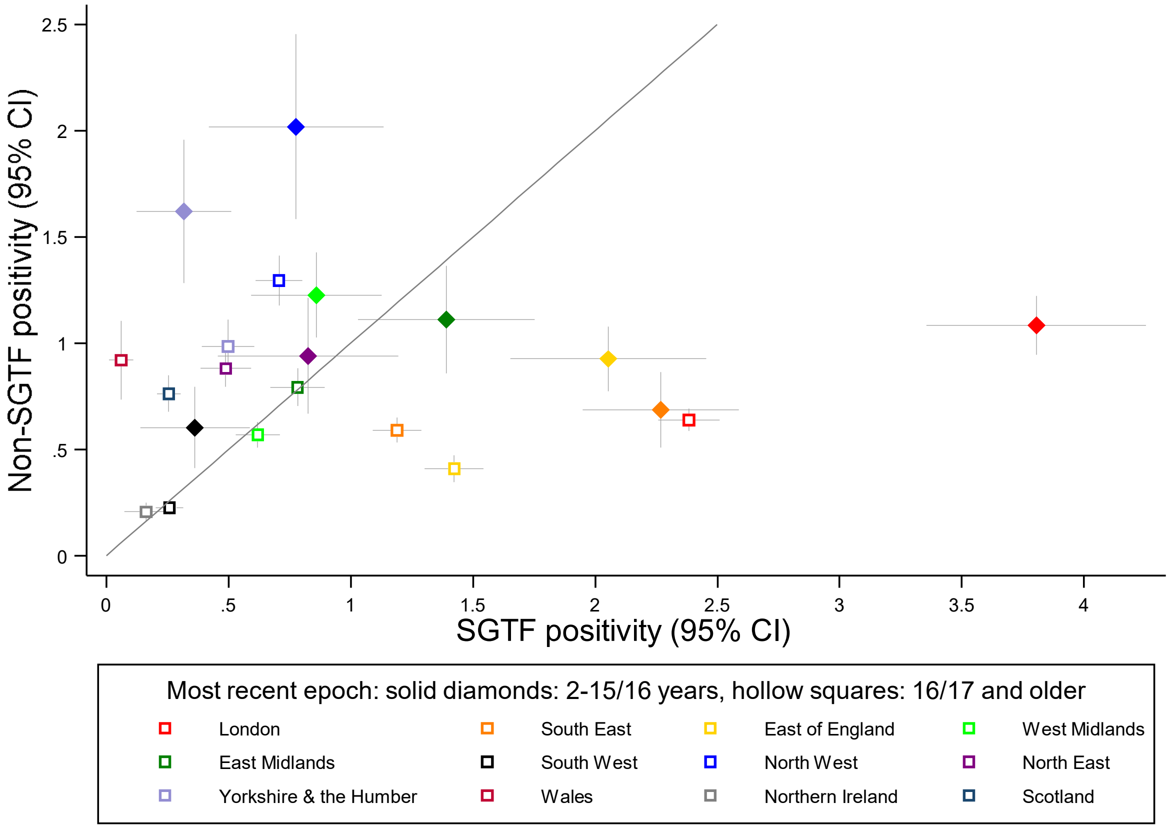

**Supplementary Figure 15 Positivity rates by age for SGTF (A) and non-SGTF (B) by English region**

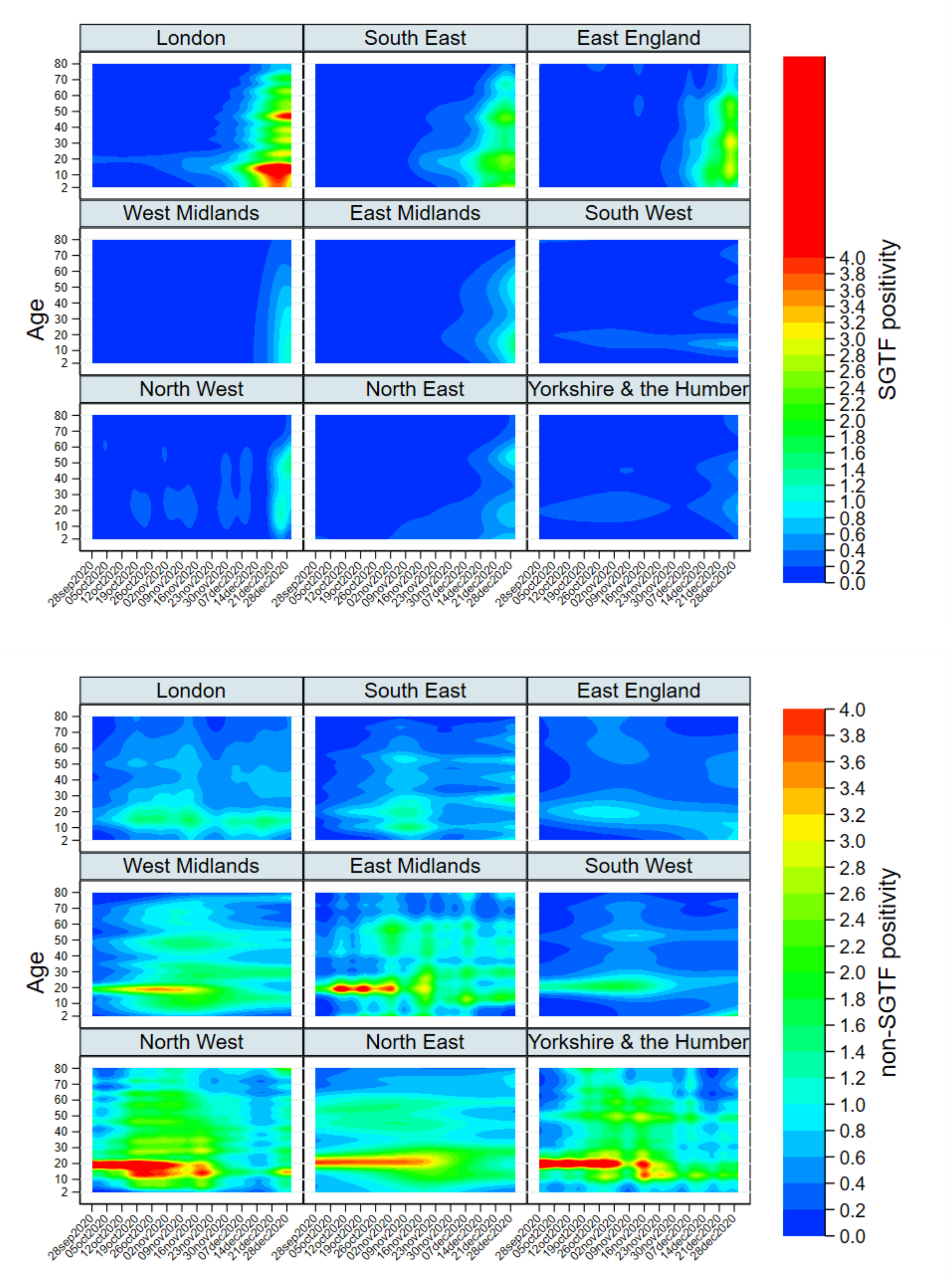

**Supplementary Table 1 Changepoints, growth rates and current predicted positivity rates from ISR models for SGTF (A) and non-SGTF positives (B)**

**Supplementary Table 1 Changepoints, growth rates and current predicted positivity rates from ISR models for SGTF (A, C) and non-SGTF positives (B, D) for those aged 2 to school year 11 (15/16 years) (A, B) and school year 12 (16/17 years) and older**

**Supplementary Table 3 Number and percentage of positives that are SGTF vs triple gene positive vs non-SGTF 21 December 2020-2 January 2021**
